# An open-label prospective observational study of antiandrogen and non-antiandrogen early pharmacological approaches in females with mild-to-moderate COVID-19. The Pre-AndroCoV Female Trial

**DOI:** 10.1101/2020.10.05.20206870

**Authors:** Flavio A. Cadegiani, Andy Goren, Carlos G. Wambier, John McCoy

## Abstract

**Background:** While COVID-19 remains largely unclear and mortality continues to raise, early effective approaches prior to complications lack, as well as researches for characterization and therapeutical potential options in actual early COVID-19. Although females seem to be less affected than females, hyperandrogenic (HA) phenotype, like polycystic ovary syndrome (PCOS), idiopathic hirsutism, congenital adrenal hyperplasia (CAH) female androgenetic alopecia (AGA), or idiopathic HA may be at higher risk due to its inherent enhanced androgenic activity. The present study aimed to evaluate the effects of any early pharmacological approach to females diagnosed with COVID-19 before seven days of symptoms, as well as investigate whether HA is an additional risk factor in this population.

**Materials and methods:** Females with symptoms for less than seven days confirmed for COVID-19 through positive real-time polymerase chain reaction (rtPCR-SARS-CoV-2) were classified and divided as non-HA, HA, and HA using spironolactone (HA-spiro) groups. Patients were questioned for baseline characteristics, 23 different diseases, 44 drug classes and vaccines, 28 different symptoms, and eight different parameters to measure COVID-19 related clinical outcomes. Treatment was then provided, including azithromycin 500mg/day for five days in all cases, associated with hydroxychloroquine 400mg/day for five days, nitazoxanide 500mg twice a day for six days, or ivermectin 0.2mg/kg/day por three days, and optionally spironolactone 100mg twice a day until cure. Patients were assessed for COVID-19 clinical course, clinical and viral duration, and disease progression.

**Results:** In total, 270 females were enrolled, including 195, 67, and eight in non-HA, HA, and HA-spiro groups, respectively. Prevailing symptoms were anosmia (71.1%), ageusia (67.0%), headache (48.1%), myalgia (37.4%), dry cough (36.3%), nasal congestion or rhinorrhea (34.1%), fatigue (33.3%), weakness (29.5%), hyporexia (27.8%), thoracic pain (24.8%), diarrhea (24.1%) and dizziness (21.5%). Earliest symptoms (days) were dizziness (*1*.*0 ± 0*.*2 day), abdominal pain* (*1*.*1 ± 0*.*3);* conjunctival hyperemia (*1*.*1 ± 0*.*5)*, nasal congestion or rhinorrhea (*1*.*2 ± 0*.*5)*, headache (*1*.*2 ± 0*.*5), dry cough* (*1*.*2 ± 0*.*5)*, myalgia (*1*.*2 ± 0*.*4)*, nauseas (*1*.*3 ± 0*.*5)* and weakness (*1*.*3 ± 0*.*5)*. Time-to-treat, positive rtPCR, and duration of symptoms with and without anosmia and ageusia were significantly lower in HA-spiro than non-HA, HA, and overall non-users. Time-to-treat was similar while all duration of symptoms and positive rtPCR-SARS-CoV-2 were significantly shorter in non-HA than HA. Spironolactone users were more likely to be asymptomatic than non-users during COVID-19. Fewer non-HA than HA females were affected by anosmia, ageusia, dry cough, fatigue, weakness and hyporexia. Ageusia, weakness and myalgia lasted shorter in non-HA than HA. None of the patients needed hospitalization or any other COVID-19 complication.

**Conclusions:** A sensitive, early detection of COVID-19 followed by a pharmaceutical approach with different drug combinations yielded irrefutable differences compared to sex-, age-, body mass index (BMI)-, and disease-matched non-treated controls in terms of clinical outcomes, ethically disallowing placebo-control randomized clinical trials in the early stage of COVID-19 due to the marked improvements. HA females presented more severe and prolonged clinical manifestations, although none progressed to worse outcomes. Spironolactone mitigated the additional risks due to HA.

## Background

COVID-19 is a multi-systemic and multi-factorial syndrome caused by the Severe Acute Respiratory Syndrome Coronavirus 2 (SARS-CoV-2). Its exact mechanisms of action are still largely unclear, and despite the massive number of infections and deaths, effective approaches before it becomes severe lack (1-4).

One of the most likely reasons to explain why we have failed to detect effective approaches is that while we are searching for molecules with antiviral activity, we are detecting COVID-19 too late, when viral infectivity no longer plays a key role in the pathophysiology at that stage, which will naturally lead to lack of efficacy from these antiviral approaches.

While we have focused the vast majority of the researches on patients after they acute lung injury and hospitalized patients, a relative shortage of researches in actual earlier stages of COVID-19, in comparison to the relevance of trying to discover effective approaches for secondary prevention, i.e., preventions of COVID-19 complications after its detection. Meanwhile. number of researches that allege to have researched in mild patients actually included hospitalized patients only, which is inherently contradictory **(5)**.

Because of the large pre-symptomatic period, asymptomatic infected subjects, prolonged incubation and viral shedding period, and unrevealed means of transmission, viral spreading remains, despite all unprecedented public policies.

Yet the most characteristic and specific symptoms have been extensively described, unspecific clinical manifestations, particularly in the first days of the disease (since anosmia and ageusia tend to appear after three a five days only), highly heterogeneous clinical presentation, and lack of good predictors of those who will further develop acute lung injury are still challenges to detect COVID-19 during the period when therapies focusing on antiviral activity may still be effective (6-9).

An additional challenge to be overcome it the persistence of policies focusing on the *sine-quo-non* presence of fever or shortness of breath to perform the diagnostic real-time Polymerase Chain Reaction (rtPCR) for SARS-CoV-2 (10,11). While these two symptoms should not be considered as signs for the presence of COVID-19, but for severe COVID-19 instead, we will fail to diagnose COVID-19 when complications are potentially avoidable. The reports on the literature claiming that fever is present in the majority of patients with COVID-19 are based on data collected from registers that require fever to diagnose COVID-19, which is *per se* a limitation for a more accurate description of COVID-19 manifestations. Reports based on diagnostic tools for COVID-19 that do not require fever show that fever may be present in as low as 10% of infected patients only (12-14).

Several different molecules demonstrated *in vitro* antiviral activity against SARS-CoV-2 and have been proposed as promising therapies for COVID-19 (13), among which the most attempted drug combinations included azithromycin in the majority of the cases, in association with hydroxychloroquine, ivermectin or nitazoxanide (13,14). However, since detection of COVID-19 is predominantly delayed due the mandatory presence of fever for its suspect, antiviral approaches will be less effective, since at this stage of the disease viral infectivity becomes less central. Accordingly, randomized clinical trials (RCT) on alleged early COVID-19 yielded conflicting results, although the majority have been exclusively performed in hospitalized patients (10-14).

We have concluded that pharmacological therapies for truly early and mild COVID-19 has not been investigated thoroughly, which precludes from conclusive findings regarding the efficacy of antiviral approaches at this stage. To evaluate potential antiviral therapies, it is critical to detect COVID-19 during the first days after its appearance, which is only feasible with more sensitive approaches its diagnosis.

Although underreported, risks for acute lung injury, thrombosis and other clinical complications in COVID-19 are also related to increased exposure, enhanced activity, and/or hypersensitivity to androgens (15-23). Overrepresentation of males in terms of complications related to COVID-19 are not fully justified by differences in age, body mass index (BMI), prevalence of comorbidities, *i*.*e*., there is an inherent risk related to male sex (15-18). This is likely explained, at least partially, by the transmembrane serine protease 2 (TMPRSS-2), a critical protein for the SARS-CoV-2 entry into the cells, that are largely and solely regulated by androgens. Among males, androgenetic alopecia (AGA) as an independent predictor of complications related to COVID-19, possibly due to a resultant of overexpression of androgen receptors (AR), due to enhanced dihydrotestosterone (DHT) levels, activity, response, or a combination between these factors, that discloses AGA as a clinical phenotype expression.

While females are underrepresented among severe COVID-19 patients, risk factors including menopause, aging, uncompensated type 2 diabetes mellitus (T2DM), hypertension and obesity seem to enhance the risk of severe COVID-19 in females more than in males. In addition, in an analogically similar manner than AGA males, females with any expression of hyperandrogenism (HA), including polycystic ovary syndrome (PCOS), idiopathic hirsutism, congenital adrenal hyperplasia (CAH) due to 21alpha-hydroxylase or 11beta-hydroxylase deficiency, female AGA, or idiopathic HA, have sufficient mechanistical plausibility to support the hypothesis that this population may be at higher risk compared to non-HA females.

In this regard, the use of antiandrogens has demonstrated promising results, as already observed for both males and females, at least when used chronically (19-22). This reinforces the role of the role of antiandrogen approaches as an additional path to improve outcomes in COVID-19. Nonetheless, similarly to the use of antiviral therapies, antiandrogens should be tested during the first stage, as it affects viral infectivity.

There were sufficient theoretical, mechanistical, observational and epidemiological observations to intuitively hypothesize that if the lack of sensitivity is to detectCOVID-19 is addressed and therefore diagnosed during the first stage, preferably before seven days of symptoms, antiviral pharmacological attempts could be then effective. At this point, it is uncertain whether this approach would be affective, which is our objective.

Together, the evaluation of sex differences, as well as differences between phenotypes within each sex, would also disclose additional information for promising approaches for specific populations.

The objectives of the present study are to perform a thorough and comprehensive clinical characterization of patients with COVID-19 detected through a highly sensitive case-detection basis, and to explore the clinical responses and outcomes to a variety of drug combinations. In addition, we aimed to detect sex-specific and androgenic phenotype-specific clinical manifestations and outcomes. This is an open-label prospective observational study performed alongside with our currently ongoing RCT (ClinicalTrials.gov Identifier: NCT04446429. Available at clinicaltrials.gov (https://clinicaltrials.gov/ct2/show/NCT04446429?term=NCT04446429&draw=2&rank=1). The present study has received approval from the Institutional Review Board (IRB) of the Ethics Committee of the National Board of Ethics Committee of the Ministry of Health, Brazil (CEP/CONEP: Parecer 4.173.074 / CAAE: 34110420.2.0000.0008).

## Materials and methods

### Subject selection

This specific study is an open label prospective observational study of the characterization and clinical outcomes of females with COVID-19 in response to specific therapeutic combinations. In order to detect cases during the earliest stages of COVID-19, we employed a highly sensitive case-detection criteria for suspect for COVID-19. We changed from the mandatory presence of severity or specific signs (shortness of breath, fever, anosmia, ageusia) to the occurrence of absolutely any atypical symptom or changes in patters of chronic symptoms, even when not supposedly related to COVID-19. Suspected females underwent rtPCR-SARS-CoV-2 (Abbott RealTime SARS-CoV-2 Assay, Abbott, USA; or Cobas SARS-CoV-2, Roche, Switzerland), and those confirmed for SARS-CoV-2 were included.

Inclusion criteria included: 1. Confirmed COVID-19 through positive rtPCR-SARS-CoV-2; 2. 18 years old and above; 3. Less than seven days of beginning of symptoms; 4. Less than 72 hours of COVID-19 confirmation (in case COVID-19 had already been confirmed); 4. Non use or use for less than 24 hours of any potential antiviral drug; and 5. No previous use of glucocorticoid in the past seven days.

### Design and methods

Parameters evaluated by the present study are depicted in Table 1. Females included in the study were actively questioned for baseline and medical characteristics, including 23 different diseases, 44 drug classes and vaccines, 28 different symptoms, in addition to the search for HA, which includes: 1. PCOS confirmed by two of three Rotterdam Criteria; 2. Previously diagnosed CAH; 3. Known hyperandrogenism, clinical- or biochemically; 4. Hirsutism; and 5. Female AGA. Females were then divided according to the presence or absence of HA (non-HA group), and in case of hyperandrogenism, use of spironolactone 100mg/day or above (HA-Spiro group) or not (HA group).

**Table 1.** Parameters evaluated for the present prospective observational study.

| Aspect | Parameter |
| --- | --- |
| **Baseline characteristics** |  |
|  | Age (y/o) |
|  | Weight (kg) |
|  | Height (m) |
|  | BMI (kg/m<sup>2</sup>) |
|  | Married (yes/no) and households (yes/no) |
| **Disease and treatment timing** |  |
|  | Time-to-treat (interval between beginning of symptoms and beginning of specific treatment) (days) |
|  | Duration of positive rtPCR SARS-CoV-2 (days) |
|  | Duration of symptoms (not including anosmia and ageusia) (days) |
|  | Duration of symptoms (including anosmia and ageusia) (days) |
| **Medical history** |  |
| Existing diseases | Hypertension |
|  | Myocardial infarction |
|  | Stroke |
|  | Chronic heart failure |
|  | Lipid disorders |
|  | Type 2 diabetes mellitus (T2DM) |
|  | Pre-diabetes |
|  | Dysglycaemia (T2DM + pre-diabetes) |
|  | Obesity |
|  | Asthma |
|  | Chronic Obstructive Pulmonary Disease (COPD) |
|  | Chronic Kidney Disease (CKD) |
|  | Liver fibrosis or cirrhosis |
|  | Major depression |
|  | Anxiety or anxiety-related disorders |
|  | Attention deficiency and hyperactive disorders (ADHD) |
|  | Insomnia |
|  | Hypothyroidism |
|  | Autoimmune disorders (any) |
|  | Current or previous non-breast non-thyroid cancer |
|  | Current or previous breast cancer |
|  | Current or previous thyroid cancer |
|  | Menopause |
|  | Endometriosis |
|  | Other diseases (any) |
| **Current medications** |  |
| Cardiovascular drugs | Beta-blocker<br>Angiotensin converter inhibitors (ACEi) (-pril)<br>Angiotensin-2 receptor blockers (ARB) (-tan)<br>Loop diuretics (furosemide)<br>Thiazide diuretics (hydrochlorothiazide (HCTZ), indapamide)<br>Calcium channel blockers (CCB) (-dipine)<br>K-sparing diuretics (spironolactone)<br>Statins (pitava-, rosuva-, atorva-, prava-, simvastatin)<br>Other lipid-lowering agents (fibrates, ezetimibe, PCSK9 inhibitors)<br>Aspirin<br>Clopidogrel<br>Warfarin<br>Xa factor inhibitors (apibaxan, rivaroxaban)<br>Direct thrombin inhibitors (dabagatran)<br>Heparins |
| Diabetes, obesity, and metabolic-related drugs | Biguanides (metformin)<br>Glucagon-like peptide 1 (GLP1) receptor analogues (GLP-1Ra) (lira-, sema-, dulaglutide; exenatide)<br>Sodium-glucose co-transporter 2 (SGLT2) inhibitors (SGLT2i) (empa-, dapa-, canagliflozin)<br>Di-peptyl peptidase 4 (DPP4) inhibitors (DPP4i) (vilda-, sita-, saxa-, linagliptin)<br>Sulfonylureas (glipizide, glimepiride, glicazide)<br>Glitazone<br>Acarbose<br>Insulin<br>Orlistat |
| Hormone and related therapies | Levothyroxine (with or without liothyronine)<br>Oral contraceptives<br>Hormonal replacement therapy (HRT) for menopause<br>Other hormonal regimes<br>Aromatase inhibitors (anastrozole; letrozole)<br>Selective estrogen receptor modulators (SERMs) |
| Central-acting drugs | Hypnotics (zolpidem, zopiclone, eszopiclone, ramelteon)<br>Selective serotonin reuptaker inhibitors (SSRIs) (fluoxetine, (des)venlafaxine, sertraline, (es)citalopram, vortioxetine, fluvoxamine)<br>Other antidepressants and humor stabilizers (bupropion, topiramate, trazodone, ami- ou nortriptiline, topiramate, oxcarbamazepine)<br>Benzodiazepines (Lora-, bromo-, dia-, clonazepam; alpra-, midazolam)<br>Atypical antipsychotics (olanzapine, quetiapine, risperidone, clozapine, aripiprazole)<br>Central nervous system (CNS) stimulants (methylfenidate, lisdexamfetamine, modafinil) |
| Other drugs |  |
| Supplements | Omega-3 (> 3g/day)<br>Vitamina D (> 1,000iu/day)<br>Zinc (> 15mg/day)<br>Vitamin C (> 500mg/day) |
|  | Multivitamins |
| **Vaccine** | BCG (lifetime)<br>Influenza (in 2020)<br>Pneumococcal 13 or 23 (since 2017) |
| **Lifestyle** | Current smoking (> 2 packs/week and > 10 pack-year)<br>Regular physical activity (> 150min/week, moderate-to-vigorous, > 3 METs, for > 1y) |
| **Clinical characterization**<br>- | Presence (yes/no)<br>Time to appearance (days)<br>Duration (days) |
| Unspecific symptoms | Fever<br>“Feverish”<br>Headache<br>Shortness of breath<br>Anosmia<br>Ageusia / hypergeusia / dysgeusia<br>Dizziness<br>Weakness<br>Fatigue<br>Hyporexia / anorexia<br>Thoracic pain<br>Lower back pain<br>Dry eyes / dry mouth / skin lesions<br>Breast pain |
| Upper respiratory tract infection-like symptoms | Nasal congestion or rhinorrhea<br>Dry cough<br>“Sinusitis” (self-reported perception)<br>“Sore throat” (self-reported perception) |
| Dengue-like symptoms | Myalgia<br>Arthralgia<br>Upper back pain<br>Conjunctival hyperemia<br>Pre-orbital pain |
| Gastrointestinal (GI) infection-like symptoms | Diarrhea<br>Nauseas<br>Vomiting<br>Abdominal or pelvic pain |
| Clinical clustering | Anosmia-Ageusia dominance; or<br>Dengue-like symptomatology / clinical presentation; or<br>URTI-like symptomatology / clinical presentation; or<br>GI infection-like symptomatology / clinical presentation; or<br>Mixed; or<br>Unspecific; or<br>Asymptomatic |
| **Treatment** |  |
| (Azithromycin 500mg/day for 05 days +) | Hydroxychloroquine 400mg/day for 05 days, or<br>Nitazoxanide 500mg BID for 06 days, or<br>Ivermectin 0.2mg/kg/day for 03 days<br>+/-<br>Spironolactone 100mg BID for 15 days, or<br>Dutasteride 0.5mg/day until cure |
| Additional treatments<br>(added according to clinical judgement) | Xa factor inhibitors<br>Warfarin<br>Enoxaparin<br>Acetylsalicylic acid (ASA)<br>Glucocorticoids (methylprednisolone, dexamethasone, prednisone, prednisolone)<br>Bromhexine<br>N-acetylcysteine<br>Colchicine<br>Vitamin C (additional dose, if already under use)<br>Zinc (additional dose, if already under use)<br>Vitamin D (additional dose, if already under use) |
| **Outcomes (Day 0 = beginning of treatment)** |  |
| WHO COVID Ordinal Outcomes<br>(Stages 1-5) | Day 0<br>Day 7<br>Day 14<br>Day 30<br>Day 60 |
| Loss of ability to everyday activities (0-100; 0 = no loss; 100 = complete inability) | Day 0<br>Day 3<br>Day 7<br>Day 14<br>Day 30 |
| Symptom severity (0-100; 0 = worst day of symptoms; 100 = no symptoms or fully recovered) | Day -7 to -4<br>Day -3 to -1<br>Day 0<br>Day 1<br>Day 2<br>Day 3<br>Day 7<br>Day 14<br>Day 21<br>Day 30<br>Day 60 |
| Thoracic CT scan (% of lungs affected) | Day 0<br>Day 7<br>Day 14<br>Day 30 |
| **Disease progression outcomes** |  |
|  | Brescia COVID-19 Respiratory Severity Scale (0-4)<br>Hospitalization<br>Intensive Care Unit (ICU)<br>Mechanical ventilation<br>Noradrenaline/dopamine<br>Death |

Clinical presentations were clustered into one of the following 1. Anosmia-Ageusia dominance; 2. Dengue-like symptomatology / clinical presentation; 3. Upper respiratory tract infection (URTI) URTI-like symptomatology / clinical presentation; 4. Gastrointestinal (GI) infection-like symptomatology / clinical presentation; 5. Mixed between clusters; 6. Unspecific presentation; or 7. Asymptomatic.

To fill criteria for each cluster, it has been required for:

1. Anosmia-Ageusia dominance: at least two of nasal congestion or rhinorrhea, dry cough, self-reported perception of “sinusitis”, or self-reported perception of “sore throat”;
2. Dengue-like clinical presentation: at least three of myalgia, arthralgia, upper back pain, conjunctival hyperemia or pre-orbital pain;
3. URTI-like clinical presentation: at least two of nasal congestion or rhinorrhea, dry cough, self-reported perception of “sinusitis”, or self-reported perception of “sore throat”;
4. GI infection-like clinical presentation: at least two of diarrhea, nauseas, vomiting, or abdominal pain;
5. Mixed between clusters: when there are symptoms to fill criteria for at least two clusters
6. Unspecific presentation: when there are only unspecific or insufficient symptoms to fill criteria for any cluster; or
7. Asymptomatic.

After characterization, drug combination including azithromycin 500mg/day for five days, with hydroxychloroquine 400mg/day for five days, nitazoxanide 500mg twice a day for six days, or ivermectin 0.2mg/kg/day for three days was then provided. The choice between hydroxychloroquine, nitazoxanide, and/or ivermectin was based on an almost-random manner, i.e., randomly, except when clinical judgement considered otherwise. In addition, spironolactone, vitamin D, vitamin C, zinc, apibaxan, rivaroxaban, enoxaparin, and glucocorticoids could have been prescribed, also according to medical judgement.

Patients were then evaluated for: 1. Time-to-appearance and duration of each symptom (number of days); 2. Time until full remission of symptoms, not including anosmia and ageusia (number of days); 3. Time until full remission of symptoms, including anosmia and ageusia (number of days); 4. Duration of positive rtPCR-SARS-CoV-2 (in number of days, where rtPCR was performed every seven days); 5. Level of clinical improvement in Days -7 to -4, -3 to -1, 0, 1, 2, 3, 7, 14, 21, 30 and 60 days, where 0 corresponds to the worst day of symptoms (scored according to the number and severity of symptoms) and 100 means asymptomatic or entirely recovered; 6. Ability to perform everyday activities in Days 0, 3, 7, 14 and 30 (0 = no loss of capacity and 100 = complete inability to perform any self-care or everyday activity); 7. WHO COVID Ordinal Outcomes Scale; 8. Brescia COVID-19 score; 9. Disease progression outcomes, including hospitalization, mechanical ventilation, and death. Full raw data is available at a data repository (https://osf.io/cm4f8/).

### Statistical analysis

Nonparametric ANOVA (Kruskal-Wallis) was performed for all parameters, regardless of the distribution normality, and *post-hoc* adjusted Dunn’s test was performed for subgroup analyses, whenever *p* <0.05. All statistical tests were performed using XLSTAT (Microsoft, USA).

## Results

Tables 6 to 15 detail characteristics and parameters of overall females and for each group (non-HA, HA and HA-spiro), and overall and pairwise comparisons. Tables 2 to 5 depict baseline and medical characteristics, Tables 6 to 8 describe COVID-19 presentation, Tables 9 and 10 show the proposed pharmacological interventions for COVID-19, and Tables 11 to 15 depict COVID-19 clinical outcomes.

### Patients’ characterization

In total, 270 females confirmed for COVID-19 were included. Of these, 195, 67, and eight were from the non-HA, HA, and HA-spiro groups. The dropout rate for clinical characterization and disease outcomes was zero.

Baseline characteristics are described in Table 2. HA females were significantly younger, shorter, and heavier than non-HA.

**Table 2.**
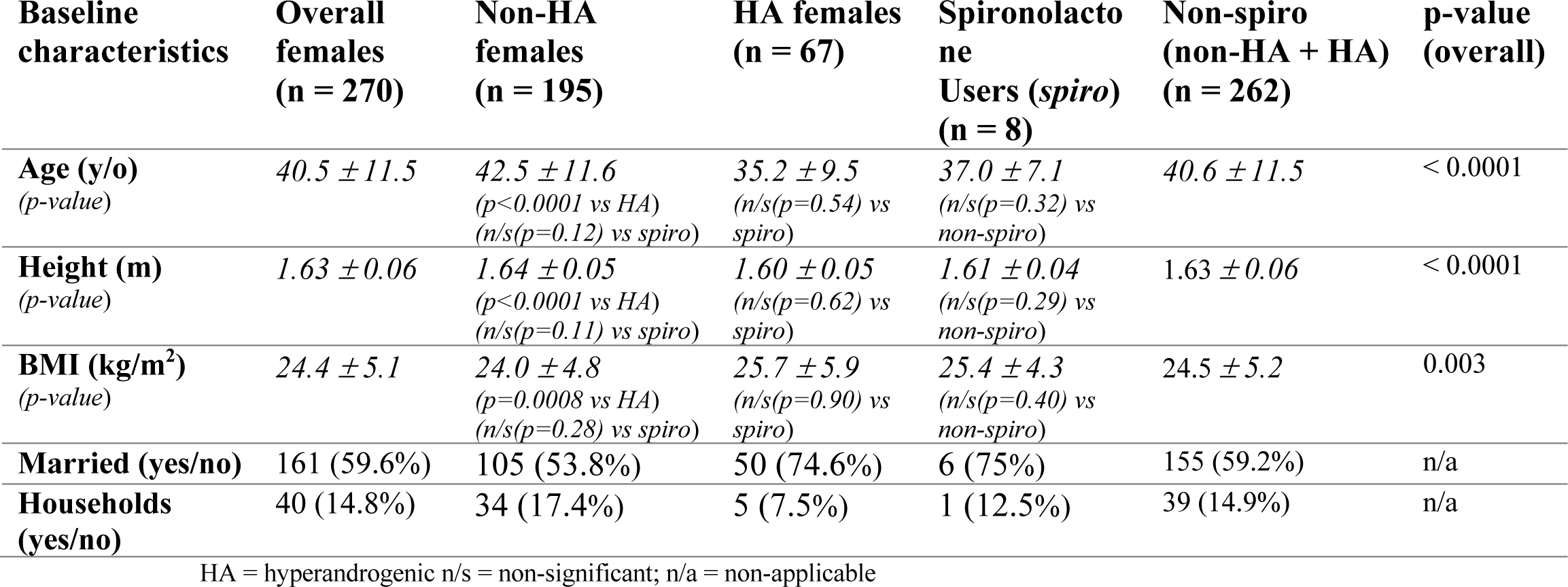
Baseline characteristics.

| **Baseline characteristics** | **Overall females (n = 270)** | **Non-HA females (n = 195)** | **HA females (n = 67)** | **Spiro lactone Users (spiro) (n = 8)** | **Non-spiro (non-HA + HA) (n = 262)** | **p-value (overall)** |
| --- | --- | --- | --- | --- | --- | --- |
| **Age (y/o)**<br>(*p-value*) | 40.5 ± 11.5 | 42.5 ± 11.6<br>(*p* < 0.0001 vs HA)<br>(*n/s* (*p* = 0.12) vs spiro) | 35.2 ± 9.5<br>(*n/s* (*p* = 0.54) vs spiro) | 37.0 ± 7.1<br>(*n/s* (*p* = 0.32) vs non-spiro) | 40.6 ± 11.5 | < 0.0001 |
| **Height (m)**<br>(*p-value*) | 1.63 ± 0.06 | 1.64 ± 0.05<br>(*p* < 0.0001 vs HA)<br>(*n/s* (*p* = 0.11) vs spiro) | 1.60 ± 0.05<br>(*n/s* (*p* = 0.62) vs spiro) | 1.61 ± 0.04<br>(*n/s* (*p* = 0.29) vs non-spiro) | 1.63 ± 0.06 | < 0.0001 |
| **BMI (kg/m<sup>2</sup>)**<br>(*p-value*) | 24.4 ± 5.1 | 24.0 ± 4.8<br>(*p* = 0.0008 vs HA)<br>(*n/s* (*p* = 0.28) vs spiro) | 25.7 ± 5.9<br>(*n/s* (*p* = 0.90) vs spiro) | 25.4 ± 4.3<br>(*n/s* (*p* = 0.40) vs non-spiro) | 24.5 ± 5.2 | 0.003 |
| **Married (yes/no)** | 161 (59.6%) | 105 (53.8%) | 50 (74.6%) | 6 (75%) | 155 (59.2%) | n/a |
| **Households (yes/no)** | 40 (14.8%) | 34 (17.4%) | 5 (7.5%) | 1 (12.5%) | 39 (14.9%) | n/a |

HA = hyperandrogenic n/s = non-significant; n/a = non-applicable

The major and prevailing diseases were present in similarly present in all groups (Table 3), while chronic kidney disease (CKD) was present in one patient, and liver fibrosis and cirrhosis, and current cancer were absent. Although HA had greater BMI than non-HA females, prevalence of obesity was similar between groups.

**Table 3.**
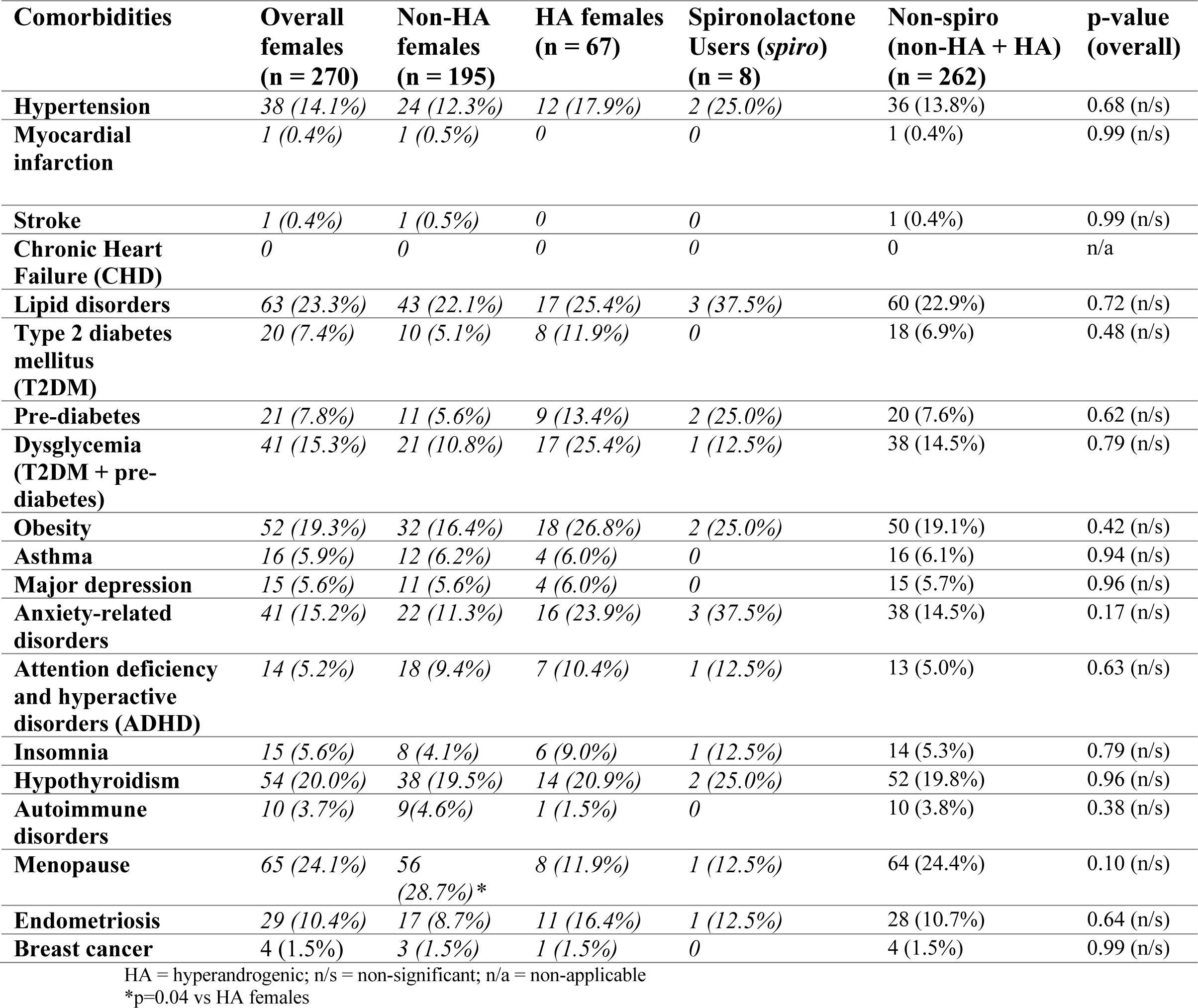
Comorbidities.

Table 4 depicts the medications used on a chronic and regular basis. Metformin and oral contraceptives were used by significantly larger number of HA than non-HA females. None of the other drugs for metabolic, cardiovascular, psychiatric or hormonal diseases disclosed differences between groups. Warfarin, direct thrombin inhibitors and heparin were not used by any patient. The percentage of participants with vaccines for BCG, influenza and pneumococcal were similar between groups, as well as practice of regular physical (Table 5).

**Table 4.** Medications used.

| **Current medications** | **Overall females (n = 270)** | **Non-HA females (n = 195)** | **HA females (n = 67)** | **Spironolactone Users (*spiro*) (n = 8)** | **Non-spiro (non-HA + HA) (n = 262)** | **p-value (overall)** |
| --- | --- | --- | --- | --- | --- | --- |
| **Beta-blocker** | 3 (1.1%) | 3 (1.5%) | 0 | 0 | 3 (1.1%) | > 0.9 (n/s) |
| **Angiotensin converter inhibitors (ACEi)** | 8 (3.0%) | 5 (2.6%) | 2 (3.0%) | 1 (12.5%) | 7 (2.7%) | > 0.9 (n/s) |
| **Angiotensin-2 receptor blockers (ARB)** | 32 (11.8%) | 19 (9.7%) | 12 (17.9%) | 1 (12.5%) | 31 (11.8%) | 0.61 (n/s) |
| **Loop diuretics** | 3 (1.1%) | 2 (1.0%) | 0 | 1 (12.5%) | 2 (0.8%) | n/a |
| **Thiazide diuretics** | 10 (3.7%) | 8 (4.1%) | 2 (3.0%) | 0 | 10 (3.8%) | > 0.9 (n/s) |
| **Calcium channel blockers (CCB)** | 16 (5.9%) | 10 (5.1%) | 6 (8.9%) | 0 | 16 (6.1%) | 0.86 (n/s) |
| **Statins** | 51 (18.9%) | 33 (16.9%) | 17 (25.4%) | 1 (12.5%) | 50 (19.1%) | 0.56 (n/s) |
| **Aspirin** | 1 (0.4%) | 1 (0.5%) | 0 | 0 | 1 (0.4%) | n/a |
| **Clopidogrel** | 1 (0.4%) | 1 (0.5%) | 0 | 0 | 1 (0.4%) | n/a |
| **Xa factor inhibitors** | 3 (1.1%) | 2 (1.0%) | 0 | 1 (12.5%) | 2 (0.8%) | n/a |
| **Metformin** | 51 (18.9%) | 26 (13.3%)* | 22 (32.8%) | 3 (37.5%) | 48 (18.3%) | 0.038 |
| **Glucagon-like peptide 1 (GLP1) receptor analogues (GLP-1Ra)** | 20 (7.4%) | 9 (4.6%) | 10 (14.9%) | 1 (12.5%) | 19 (7.2%) | 0.44 (n/s) |
| **Sodium-glucose co-transporter 2 (SGLT2) inhibitors (SGLT2i)** | 30 (11.1%) | 16 (8.2%) | 12 (17.9%) | 2 (25.0%) | 28 (10.7%) | 0.39 (n/s) |
| **Di-peptyl peptidase 4 (DPP4) inhibitors (DPP4i)** | 6 (2.2%) | 4 (2.0%) | 1 (1.5%) | 1 (12.5%) | 5 (1.9%) | 0.98 (n/s) |
| **Sulfonylureas** | 0 | 0 | 0 | 0 | 0 | n/a |
| **Pioglitazone** | 0 | 0 | 0 | 0 | 0 | n/a |
| **Insulin** | 1 (0.4%) | 1 (0.5%) | 0 | 0 | 1 (0.4%) | n/a |
| **Orlistat** | 12 (4.4%) | 2 (1.0%) | 10 (14.9%) | 0 | 12 (4.6%) | 0.23 (n/s) |
| **Levothyroxine** | 54 (20.0%) | 38 (19.5%) | 14 (20.9%) | 2 (25.0%) | 52 (19.8%) | 0.84 (n/s) |
| **Liothyronine** | 3 (1.1%) | 3 (1.5%) | 0 | 0 | 3 (1.1%) | 0.77 (n/s) |
| **Estradiol (E; no P)** | 7 (2.6%) | 6 (3.1%) | 1 (1.5%) | 0 | 7 (2.7%) | 0.45 (n/s) |
| **Progesterone (P; no E)** | 0 | 0 | 0 | 0 | 0 | n/a |
| **Combined therapy (E+P) (for menopause)** | 29 (10.7%) | 24 (12.3%) | 5 (7.5%) | 0 | 29 (11.1%) | 0.73 (n/s) |
| **Oral contraceptives** | 36 (13.3%) | 17 (8.7%)* | 19 (28.4%) | 0 | 36 (13.7%) | 0.045 |
| **Other hormonal regimens** | 0 | 0 | 0 | 0 | 0 | n/a |
| **Aromatase inhibitors (anastrozole; letrozole) or Selective estrogen receptor modulators (SERMs)** | 3 (1.1%) | 1 (0.5%) | 2 (3.0%) | 0 | 3 (1.1%) | 0.85 (n/s) |
| **Hypnotics** | 19 (7.0%) | 10 (5.1%) | 8 (11.9%) | 1 (12.5%) | 18 (6.9%) | 0.68 (n/s) |
| **Selective serotonin reuptaker inhibitors (SSRIs)** | 43 (15.9%) | 27 (13.8%) | 13 (19.4%) | 3 (37.5%) | 40 (15.3%) | 0.77 (n/s) |
| **Other antidepressants and humor stabilizers** | 27 (10.0%) | 15 (7.7%) | 11 (16.4%) | 1 (12.5%) | 26 (10.0%) | 0.57 (n/s) |
| **Benzodiazepines** | 3 (1.1%) | 2 (1.0%) | 1 (1.4%) | 0 | 3 (1.1%) | 0.99 (n/s) |
| **Atypical antipsychotics** | 7 (2.6%) | 4 (2.0%) | 3 (4.5%) | 0 | 7 (2.7%) | 0.91 (n/s) |
| **Central nervous system (CNS) stimulants** | 18 (6.7%) | 8 (4.1%) | 8 (11.9%) | 2 (25.0%) | 16 (6.1%) | 0.42 (n/s) |
| **Finasteride** | 4 (1.5%) | 0 | 4 (6.0%) | 0 | 4 (1.5%) | 0.76 (n/s) |
| **Oral minoxidil** | 10 (3.7%) | 4 (2.0%) | 6 (8.9%) | 0 | 10 (3.8%) | 0.69 (n/s) |
| **Omega 3** | 3 (1.1%) | 2 (1.0%) | 1 (1.5%) | 0 | 3 (1.1%) | 0.99 (n/s) |
| **Vitamin D** | 33 (11.1%) | 21 (10.8%) | 9 (13.4%) | 3 (37.5%) | 30 (11.4%) | 0.44 (n/s) |
| **Zinc** | 20 (7.4%) | 12 (6.1%) | 6 (8.9%) | 2 (25.0%) | 18 (6.9%) | 0.78 (n/s) |
| **Biotin** | 16 (5.9%) | 9 (4.6%) | 7 (10.4%) | 0 | 16 (6.1%) | 0.61 (n/s) |
| **Vitamin C** | 28 (10.4%) | 18 (9.2%) | 8 (11.9%) | 2 (25.0%) | 26 (9.9%) | 0.63 (n/s) |

HA = hyperandrogenic; n/s = non-significant; n/a = non-applicable
\*p=0.04 vs HA females

**Table 5.** Vaccines and lifestyle.

| **Vaccine and lifestyle** | **Overall females (n = 270)** | **Non-HA females (n = 195)** | **HA females (n = 67)** | **Spiroinolactone users (*spiro*) (n = 8)** | **Non-spiro (non-HA + HA) (n = 262)** | **p-value (overall)** |
| --- | --- | --- | --- | --- | --- | --- |
| **Vaccine - BCG** | 270 (100%) | 195 (100%) | 67 (100%) | 8 (100%) | 262 (100%) | 1.0 (n/s) |
| **Vaccine – Influenza (2020)** | 46 (17.0%) | 34 (17.4%) | 11 (15.5%) | 7 (13.5%) | 43 (16.4%) | 0.95 (n/s) |
| **Vaccine – Pneumococcal (since 2017)** | 37 (13.7%) | 27 (13.8%) | 10 (14.1%) | 6 (11.5%) | 35 (13.5%) | 0.97 (n/s) |
| **Current smoking** | 2 (0.7%) | 2 (1.0%) | 0 | 0 | 2 (0.8%) | 0.98 (n/s) |
| **Regular physical activity** | 76 (28.1%) | 54 (27.7%) | 20 (28.2%) | 2 (25.0%) | 71 (27.1%) | 0.99 (n/s) |

HA = hyperandrogenic; n/s = non-significant

### COVID-19 clinical presentation

Table 6 describes COVID-19 clusters of clinical presentation. URTI-like syndrome was statistically more prevalent in HA than non-HA, while anosmia-ageusia predominance, dengue fever-like, GI infection-like, mixed and unspecific symptomatology were similar between groups. Spironolactone users were more likely to be asymptomatic than non-users during COVID-19.

**Table 6.** Clinical clustering.

| Clinical clustering | Overall females (n = 270) | Non-HA females (n = 195) | HA females (n = 67) | Spironolactone Users (*spiro*) (n = 8) | Non-spiro (non-HA + HA) (n = 262) | p-value (overall) |
| --- | --- | --- | --- | --- | --- | --- |
| **Anosmia-Ageusia dominance**<br>(*p*-value) | 43 (15.9%) | 34 (17.4%)<br>(*n/s* (*p*=0.50) vs HA)<br>(*n/s* (*p*=0.83) vs *spiro*) | 8 (11.9%)<br>(*n/s*(*p*=0.98) vs *spiro*) | 1 (12.5%)<br>(*n/s* (*p*=0.86) vs *no-spiro*) | 42 (16.0%) | 0.79 (*n/s*) |
| **Dengue fever-like**<br>(*p*-value) | 77 (28.5%) | 65 (33.3%)<br>(*n/s* (*p*=0.67) vs HA)<br>(*n/s* (*p*=0.32) vs *spiro*) | 20 (29.8%)<br>(*n/s*(*p*=0.43) vs *spiro*) | 1 (12.5%)<br>(*n/s* (*p*=0.34) vs *no-spiro*) | 85 (32.4%) | 0.58 (*n/s*) |
| **URTI-like**<br>(*p*-value) | 87 (32.2%) | 53 (27.2%)<br>(*p*=0.012) vs HA)<br>(*n/s* (*p*=0.92) vs *spiro*) | 32 (47.8%)<br>(*n/s*(*p*=0.29) vs *spiro*) | 2 (25.0%)<br>(*n/s* (*p*=0.72) vs *no-spiro*) | 85 (32.4%) | 0.04 |
| **GI infection-like**<br>(*p*-value) | 33 (12.2%) | 30 (15.4%)<br>(*n/s* (*p*=0.97) vs HA)<br>(*n/s* (*p*=0.85) vs *spiro*) | 11 (16.4%)<br>(*n/s*(*p*=0.89) vs *spiro*) | 1 (12.5%)<br>(*n/s* (*p*=0.88) vs *no-spiro*) | 41 (15.6%) | 0.98 (*n/s*) |
| **Mixed**<br>(*p*-value) | 35 (12.6%) | 22 (11.3%)<br>(*n/s* (*p*=0.72) vs HA)<br>(*n/s* (*p*=0.62) vs *spiro*) | 12 (17.9%)<br>(*n/s*(*p*=0.75) vs *spiro*) | 0<br>(*n/s* (*p*=0.53) vs *no-spiro*) | 34 (13.0%) | 0.59 (*n/s*) |
| **Unspecific**<br>(*p*-value) | 65 (24.1%) | 49 (25.1%)<br>(*n/s* (*p*=0.88) vs HA)<br>(*n/s* (*p*=0.23) vs *spiro*) | 16 (23.9%)<br>(*n/s*(*p*=0.27) vs *spiro*) | 0<br>(*n/s* (*p*=0.23) vs *no-spiro*) | 65 (24.8%) | 0.48 (*n/s*) |
| **Asymptomatic**<br>(*p*-value) | 27 (10.0%) | 21 (10.8%)<br>(*n/s* (*p*=0.34) vs HA)<br>(*n/s* (*p*=0.06) vs *spiro*) | 2 (3.0%)<br>(*p*=0.03) vs *spiro*) | 4 (50.0%)<br>(*p*=0.047) vs *no-spiro*) | 23 (8.8%) | 0.089 (*n/s*) |

URTI = Upper respiratory tract infection; GI = Gastrointestinal; HA = hyperandrogenic; *n/s* = non-significant

Table 7 describes the percentage of females presenting each symptom, its average duration, and time-to-appearance. Prevailing symptoms include anosmia (71.1%) and ageusia (67.0%) and headache (48.1%). Symptoms present in 20% to 40% of COVID-19 females include myalgia (37.4%), dry cough (36.3%), fever or “feverish” (34.1%), nasal congestion or rhinorrhea (34.1%), fatigue (33.3%), weakness (29.5%), hyporexia (27.8%), thoracic pain (24.8%), diarrhea (24.1%) and dizziness (21.5%). Symptoms present in fewer than 20% of patients include “sore throat” (15.9%), pre-orbital pain (12.6%), arthralgia (10.4%), conjunctival hyperemia (8.1%), nauseas (8.1%), abdominal pain (7.8%), upper back pain (7.7%), “sinusitis” (6.7%), shortness of breath (5.9%), lower back pain (5.2%), and pre-orbital pain (3.5%), dry eyes (2.2%) and dry mouth (1.1%).

**Table 7.** Clinical manifestations in COVID-19: presence (%), time-to-appearance and duration.

| Clinical manifestations | Overall females (n = 270) | Non-HA females (n = 195) | HA females (n = 67) | Spironolactone Users (*spiro*) (n = 8) | Non-spiro (non-HA + HA) (n = 262) | p-value (overall) |
| --- | --- | --- | --- | --- | --- | --- |
| **Fever** |  |  |  |  |  |  |
| Presence (%)<br>(*p*-value) | 31 (11.5%) | 19 (9.7%)<br>(*n/s* (*p*=0.32) vs HA)<br>(*n/s* (*p*=0.64) vs *spiro*) | 12 (17.9%)<br>(*n/s*(*p*=0.41) vs *spiro*) | 0<br>(*n/s*(*p*=0.57) vs *no-spiro*) | 31 (11.8%) | 0.52 |
| Time to appearance (days)<br>(*p*-value) | 1.5 ± 0.6 | 1.7 ± 0.6<br>(*n/s* (*p*=0.19) vs HA)<br>(*n/a* vs *spiro*) | 1.3 ± 0.5<br>(*n/a* vs *spiro*) | -<br>(*n/a*) | 1.5 ± 0.6 | n/a |
| Duration (days)<br>(*p*-value) | 2.3 ± 0.8 | 2.4 ± 0.7<br>(*n/s* (*p*=0.48) vs HA)<br>(*n/a* vs *spiro*) | 2.2 ± 1.0<br>(*n/a* vs *spiro*) | -<br>(*n/a*) | 2.2 ± 0.8 | n/a |
| **“Feverish”** |  |  |  |  |  |  |
| Presence (%)<br>(*p*-value) | 61 (22.6%) | 39 (20.0%)<br>(*n/s* (*p*=0.12) vs HA)<br>(*n/s* (*p*=0.38) vs *spiro*) | 22 (32.8%)<br>(*n/s*(*p*=0.13) vs *spiro*) | 0<br>(*n/s*(*p*=0.26) vs *no-spiro*) | 61 (23.3%) | 0.16 |
| Time to appearance (days)<br>(*p*-value) | 1.5 ± 0.6 | 1.4 ± 0.5<br>(*n/s* (*p*=0.87) vs HA)<br>(*n/a* vs *spiro*) | 1.4 ± 0.7<br>(*n/a* vs *spiro*) | 0<br>(*n/a* vs *no-spiro*) | 1.4 ± 0.6 | n/a |
| Duration (days)<br>(*p*-value) | 2.4 ± 1.2 | 2.4 ± 1.0<br>(*n/s* (*p*=0.76) vs HA)<br>(*n/a* vs *spiro*) | 2.4 ± 1.4<br>(*n/a* vs *spiro*) | 0<br>(*n/a* vs *no-spiro*) | 2.4 ± 1.2 | n/a |
| **Nasal congestion or rhinorrhea** |  |  |  |  |  |  |
| Presence (%)<br>(*p*-value) | 92 (34.1%) | 63 (32.3%)<br>(*n/s* (*p*=0.33) vs HA)<br>(*n/s* (*p*=0.73) vs *spiro*) | 27 (40.3%)<br>(*n/s*(*p*=0.48) vs *spiro*) | 2 (25.0%)<br>(*n/s*(*p*=0.65) vs *no-spiro*) | 90 (34.5%) | 0.56 |
| Time to appearance (days)<br>(*p*-value) | 1.2 ± 0.5 | 1.2 ± 0.5<br>(*n/s* (*p*=0.69) vs HA)<br>(*n/a* vs *spiro*) | 1.2 ± 0.4<br>(*n/a* vs *spiro*) | 1.0 ± 0.0 (1;1)<br>(*n/a* vs *no-spiro*) | 1.2 ± 0.5 | n/a |
| Duration (days)<br>(*p*-value) | 3.6 ± 1.8 | 3.8 ± 1.8<br>(*n/s* (*p*=0.72) vs HA)<br>(*n/a* vs *spiro*) | 3.3 ± 1.5<br>(*n/a* vs *spiro*) | 1.5 ± 0.5 (1;2)<br>(*n/a* vs *no-spiro*) | 3.6 ± 1.8 | n/a |
| **Headache** |  |  |  |  |  |  |
| Presence (%)<br>(*p*-value) | 130 (48.1%) | 89 (46.6%)<br>(*n/s* (*p*=0.12) vs HA)<br>(*n/s* (*p*=0.33) vs *spiro*) | 39 (58.2%)<br>(*n/s*(*p*=0.13) vs *spiro*) | 2 (25.0%)<br>(*n/s*(*p*=0.15) vs *no-spiro*) | 128 (48.8%) | 0.16 |
| Time to appearance (days)<br>(*p*-value) | 1.2 ± 0.5 | 1.1 ± 0.3<br>(*n/s* (*p*=0.10) vs HA)<br>(*n/a* vs *spiro*) | 1.4 ± 0.7<br>(*n/a* vs *spiro*) | 1.5 ± 0.5 (1;2)<br>(*n/a* vs *no-spiro*) | 1.2 ± 0.5 | n/a |
| Duration (days) | 5.7 ± 3.5 | 5.6 ± 3.5 | 6.0 ± 3.6 | 2.0 ± 1.0 (1;3) | 5.8 ± 3.5 | n/a |

| *(p-value)* |  | *(n/s (p=0.63) vs HA)*<br>*(n/a vs spiro)* | *(n/a vs spiro)* | *(n/a vs no-spiro)* |  |  |
| --- | --- | --- | --- | --- | --- | --- |
| **Shortness of breath** |  |  |  |  |  |  |
| Presence (%)<br>*(p-value)* | 16 (5.9%) | 10 (5.1%)<br>*(n/s (p=0.72) vs HA)*<br>*(n/s (p=0.64) vs spiro)* | 6 (8.9%)<br>*(n/s(p=0.68) vs spiro)* | 0<br>*((n/s(p=0.79) vs no-spiro)* | 16 (6.1%) | 0.85 |
| Time to appearance (days)<br>*(p-value)* | 3.8 ± 1.2 | 3.5 ± 1.4<br>*(n/s (p=0.28) vs HA)*<br>*(n/a vs no-spiro)* | 4.2 ± 0.7<br>*(n/a vs spiro)* | 0<br>*(n/a vs no-spiro)* | 3.7 ± 1.2 | n/a |
| Duration (days)<br>*(p-value)* | 2.5 ± 1.6 | 2.4 ± 1.9<br>*(n/s (p=0.72) vs HA)*<br>*(n/a vs spiro)* | 2.7 ± 0.7<br>*(n/a vs spiro)* | 0<br>*(n/a vs no-spiro)* | 2.5 ± 1.6 | n/a |
| **Anosmia** |  |  |  |  |  |  |
| Presence (%)<br>*(p-value)* | 192 (71.1%) | 133 (68.2%)<br>*(p = 0.025 vs HA)*<br>*(p = 0.0008 vs spiro)* | 58 (86.6%)<br>*(p = 0.0007 vs AGA-5ARi)* | 1 (12.5%)<br>*(p = 0.0036 vs no-5ARi)* | 191 (72.9%) | 0.001 |
| Time to appearance (days)<br>*(p-value)* | 3.4 ± 1.3 | 3.2 ± 1.2<br>*(p=0.0009 vs HA)*<br>*(n/a vs spiro)* | 3.9 ± 1.2<br>*(n/a vs spiro)* | 3<br>*(n/a vs no-spiro)* | 3.4 ± 1.3 | < 0.0001 |
| Duration (days)<br>*(p-value)* | 7.9 ± 6.2 | 7.8 ± 6.2<br>*(n/s(p=0.63) vs HA)*<br>*(n/a vs spiro)* | 8.1 ± 6.0<br>*(n/a vs spiro)* | 3<br>*(n/a vs no-spiro)* | 7.9 ± 6.2 | < 0.0001 |
| **Ageusia** |  |  |  |  |  |  |
| Presence (%)<br>*(p-value)* | 181 (67.0%) | 124 (63.6%)<br>*(p=0.015 vs HA)*<br>*(p=0.014 vs spiro)* | 56 (83.6%)<br>*(p=0.001 vs spiro)* | 1 (12.5%)<br>*(p=0.007 vs no-5ARi)* | 180 (69.7%) | 0.0013 |
| Time to appearance (days)<br>*(p-value)* | 3.4 ± 1.3 | 3.3 ± 1.4<br>*(p=0.011 vs HA)*<br>*(n/a vs spiro)* | 3.8 ± 1.2<br>*(n/a vs spiro)* | 1<br>*(n/a vs no-spiro)* | 3.5 ± 1.3 | n/a |
| Duration (days)<br>*(p-value)* | 7.0 ± 5.7 | 6.7 ± 5.6<br>*(p=0.045 vs HA)*<br>*(n/a vs spiro)* | 7.9 ± 5.7.6<br>*(n/a vs spiro)* | 3<br>*(n/a vs no-spiro)* | 7.1 ± 5.7 | n/a |
| **Dry cough** |  |  |  |  |  |  |
| Presence (%)<br>*(p-value)* | 98 (36.3%) | 63 (32.3%)<br>*(p=0.039 vs HA)*<br>*(n/s(p=0.73). vs spiro)* | 33 (49.2%)<br>*(n/s (p=0.26) vs spiro)* | 2 (25.0%)<br>*(n/s (p=0.56) vs no-spiro)* | 96 (36.6%) | 0.10 (n/s) |
| Time to appearance (days)<br>*(p-value)* | 1.2 ± 0.5 | 1.2 ± 0.5<br>*(n/s (p=0.99) vs HA)*<br>*(n/a vs spiro)* | 1.2 ± 0.5<br>*(n/a vs spiro)* | 1.0 ± 0.0 (1;1)<br>*(n/a vs no-spiro)* | 1.2 ± 0.5 | n/a |
| Duration (days)<br>*(p-value)* | 5.4 ± 3.5 | 5.5 ± 3.6<br>*(n/s (p=0.80) vs HA)*<br>*(n/a vs spiro)* | 5.5 ± 3.2<br>*(n/a vs spiro)* | 2.5 ± 0.5 (2;3)<br>*(n/a vs no-spiro)* | 5.5 ± 3.5 | n/a |
| **“Sinusitis”** |  |  |  |  |  |  |
| Presence (%)<br>*(p-value)* | 18 (6.7%) | 18 (9.2%)<br>*(n/s(p=0.83) vs HA)*<br>*(n/s(p=0.66) vs spiro)* | 0<br>*(n/s(p=0.73) vs spiro)* | 0<br>*(n/s(p=0.67) vs no-spiro)* | 18 (6.9%) | 0.89 (n/s) |
| Time to appearance (days)<br>*(p-value)* | 1.3 ± 0.5 | 1.3 ± 0.5<br>*(n/s(p=0.82) vs HA)*<br>*(n/a vs spiro)* | 0<br>*(n/a vs spiro)* | 0<br>*(n/a vs no-spiro)* | 1.3 ± 0.5 | n/a |
| Duration (days)<br>*(p-value)* | 6.0 ± 2.6 | 6.0 ± 2.8<br>*(n/s(p=0.68) vs HA)*<br>*(n/a vs spiro)* | 0<br>*(n/a vs spiro)* | 0<br>*(n/a vs no-spiro)* | 6.0 ± 2.6 | n/a |
| **“Sore throat”** |  |  |  |  |  |  |
| Presence (%)<br>*(p-value)* | 43 (15.9%) | 25 (12.8%)<br>*(n/s (p=0.086) vs HA)*<br>*(n/s(p=0.54) vs spiro)* | 18 (26.9%)<br>*(n/s(p=0.22) vs spiro)* | 0<br>*(n/s(p=0.43) vs no-spiro)* | 43 (16.4%) | 0.17 (n/s) |
| Time to appearance (days) | 1.4 ± 0.7 | 1.4 ± 0.6<br>*(n/s (p=0.87) vs HA)* | 1.4 ± 1.2<br>*(n/a vs spiro)* | 0<br>*(n/a vs no-spiro)* | 1.4 ± 0.7 | n/a |
| *(p-value)* |  | *(n/a vs spiro)* |  |  |  |  |
| Duration (days)<br>*(p-value)* | 5.6 ± 2.0 | 5.9 ± 2.1<br>*((n/s (p=0.41) vs HA)*<br>*(n/a vs spiro)* | 5.3 ± 1.7<br>*(n/a vs spiro)* | 0<br>*(n/a vs no-spiro)* | 5.6 ± 2.0 | n/a |
| **Dizziness** |  |  |  |  |  |  |
| Presence (%)<br>*(p-value)* | 58 (21.5%) | 40 (20.5%)<br>*(n/s (p=0.44) vs HA)*<br>*(n/s(p=0.33) vs spiro)* | 18 (26.9%)<br>*(n/s(p=0.22) vs spiro)* | 0<br>*(n/s(p=0.29) vs no-spiro)* | 58 (22.1%) | 0.42 (n/s) |
| Time to appearance (days)<br>*(p-value)* | 1.0 ± 0.2 | 1.0 ± 0.0<br>*(n/s (p=0.50) vs HA)*<br>*(n/a vs spiro)* | 1.1 ± 0.3<br>*(n/a vs spiro)* | 0<br>*(n/a vs no-spiro)* | 1.0 ± 0.2 | n/a |
| Duration (days)<br>*(p-value)* | 1.9 ± 1.0 | 1.8 ± 0.9<br>*(n/s (p=0.86) vs HA)*<br>*(n/a vs spiro)* | 1.9 ± 1.1<br>*(n/a vs spiro)* | 0<br>*(n/a vs no-spiro)* | 1.9 ± 1.0 | n/a |
| **Fatigue** |  |  |  |  |  |  |
| Presence (%)<br>*(p-value)* | 90 (33.3%) | 54 (27.7%)<br>*(p=0.0015 vs HA).*<br>*(n/s(p=0.18) vs spiro)* | 36 (53.7%)<br>*(p=0.013 vs spiro)* | 0<br>*((n/s(p=0.098) vs no-spiro)* | 90 (34.3%) | 0.0016 |
| Time to appearance (days)<br>*(p-value)* | 1.4 ± 0.7 | 1.4 ± 0.6<br>*((n/s(p=0.74) vs HA)*<br>*(n/a vs spiro)* | 1.5 ± 0.7<br>*(n/a vs spiro)* | 0<br>*(n/a vs no-spiro)* | 1.4 ± 0.7 | n/a |
| Duration (days)<br>*(p-value)* | 7.2 ± 4.9 | 6.8 ± 4.5<br>*((n/s(p=0.35) vs HA)*<br>*(n/a vs spiro)* | 7.7 ± 5.4<br>*(n/a vs spiro)* | 0<br>*(n/a vs no-spiro)* | 7.2 ± 4.9 | n/a |
| **Weakness** |  |  |  |  |  |  |
| Presence (%)<br>*(p-value)* | 74 (29.5%) | 47 (24.1%)<br>*(p=0.048 vs HA)*<br>*(n/s(p=0.25) vs vs spiro)* | 27 (40.3%)<br>*(n/s(p=0.063) vs vs spiro)* | 0<br>*(n/s(p=0.17) vs vs no-spiro)* | 74 (28.2%) | 0.056 (n/s) |
| Time to appearance (days)<br>*(p-value)* | 1.3 ± 0.5 | 1.3 ± 0.5<br>*(n/s(p=0.93) vs HA)*<br>*(n/s(p=0.056) vs spiro)* | 1.3 ± 0.5<br>*(n/a vs spiro)* | 0<br>*(n/a vs no-spiro)* | 1.3 ± 0.5 | n/a |
| Duration (days)<br>*(p-value)* | 2.7 ± 1.4 | 2.4 ± 1.1<br>*(p=0.031 vs HA)*<br>*(n/a vs spiro)* | 3.2 ± 1.7<br>*(n/a vs spiro)* | 0<br>*(n/a vs no-spiro)* | 2.7 ± 1.4 | n/a |
| **Myalgia** |  |  |  |  |  |  |
| Presence (%)<br>*(p-value)* | 101 (37.4%) | 73 (37.4%)<br>*(n/s (0.59) vs HA)*<br>*(n/s (p=0.073) vs spiro)* | 28 (41.8%)<br>*(n/s (p=0.054) vs spiro)* | 0<br>*(n/s (p=0.063) vs no-spiro)* | 101 (38.4%) | 0.15 (n/s) |
| Time to appearance (days)<br>*(p-value)* | 1.2 ± 0.4 | 1.2 ± 0.4<br>*(n/s (0.25) vs HA)*<br>*(n/a vs spiro)* | 1.3 ± 0.5<br>*(n/a vs spiro)* | 0<br>*(n/a vs vs no-spiro)* | 1.2 ± 0.4 | n/a |
| Duration (days)<br>*(p-value)* | 3.3 ± 1.4 | 3.1 ± 1.4<br>*(p = 0.037 vs HA)*<br>*(n/a vs spiro)* | 3.7 ± 1.4<br>*(n/a vs spiro)* | 0<br>*(n/a vs no-spiro)* | 3.3 ± 1.4 | n/a |
| **Arthralgia** |  |  |  |  |  |  |
| Presence (%)<br>*(p-value)* | 28 (10.4%) | 22 (11.3%)<br>*(n/s (p = 0.78) vs HA)*<br>*(n/s (p = 0.59) vs spiro)* | 6 (8.9%)<br>*(n/s (p = 0.68) vs spiro)* | 0<br>*(n/s (p = 0.61) vs no-spiro)* | 28 (10.7%) | 0.84 (n/s) |
| Time to appearance (days)<br>*(p-value)* | 1.5 ± 0.6 | 1.4 ± 0.6<br>*(n/s (p = 0.74) vs HA)*<br>*(n/a vs spiro)* | 1.5 ± 0.5<br>*(n/a vs spiro)* | 0<br>*(n/a vs no-spiro)* | 1.5 ± 0.6 | n/a |
| Duration (days)<br>*(p-value)* | 3.0 ± 2.4 | 3.1 ± 2.4<br>*(n/s (p = 0.67) vs HA)*<br>*(n/a vs spiro)* | 2.7 ± 2.0<br>*(n/a vs spiro)* | 0<br>*(n/a vs no-spiro)* | 3.0 ± 2.4 | n/a |
| **Hyporexia** |  |  |  |  |  |  |
| Presence (%) | 75 (27.8%) | 45 (23.1%)<br>*(p=0.008 vs HA)* | 30 (44.8%)<br>*(p=0.039 vs spiro)* | 0<br>*(n/s(p=0.17) vs no-spiro)* | 75 (28.6%) | 0.011 |
| *(p-value)* |  | *(n/s(p=0.27) vs spiro)* |  |  |  |  |
| Time to appearance (days) | 1.3 ± 0.5 | 1.2 ± 0.5<br>*(n/s(p=0.17) vs HA)*<br>*(n/a vs spiro)* | 1.4 ± 0.6<br>*(n/a vs spiro)* | 0<br>*(n/a vs no-spiro)* | 1.3 ± 0.5 | n/a |
| *(p-value)* |  |  |  |  |  |  |
| Duration (days) | 4.8 ± 2.8 | 4.9 ± 3.1<br>*(n/s(p=0.99) vs HA)*<br>*(n/a vs spiro)* | 4.6 ± 2.2<br>*(n/a vs spiro)* | 0<br>*(n/a vs no-spiro)* | 4.8 ± 2.8 | n/a |
| *(p-value)* |  |  |  |  |  |  |
| Thoracic pain |  |  |  |  |  |  |
| Presence (%) | 67 (24.8%) | 43 (22.0%)<br>*(n/s(p=0.093) vs HA)*<br>*(n/s(p=0.099) vs spiro)* | 24 (35.8%)<br>*(n/s(p=0.22) vs spiro)* | 0<br>*(n/s(p=0.24) vs no-spiro)* | 67 (25.6%) | 0.11 (n/s) |
| *(p-value)* |  |  |  |  |  |  |
| Time to appearance (days) | 2.5 ± 1.0 | 2.6 ± 0.9<br>*(n/s(p=0.61) vs HA)*<br>*(n/a vs spiro)* | 2.5 ± 1.1<br>*(n/a vs spiro)* | 0<br>*(n/a vs no-spiro)* | 2.6 ± 1.0 | n/a |
| *(p-value)* |  |  |  |  |  |  |
| Duration (days) | 4.7 ± 2.5 | 4.5 ± 2.4<br>*(n/s(p=0.31) vs HA)*<br>*(n/a vs spiro)* | 5.1 ± 2.7<br>*(n/a vs spiro)* | 0<br>*(n/a vs no-spiro)* | 4.7 ± 2.5 | n/a |
| *(p-value)* |  |  |  |  |  |  |
| Upper back pain |  |  |  |  |  |  |
| Presence (%) | 28 (10.4%) | 15 (7.7%)<br>*(n/s(p=0.15) vs HA)*<br>*(n/s (p=0.71) vs spiro)* | 13 (19.4%)<br>*(n/s (p=0.37 vs spiro)* | 0<br>*(n/s (p=0.61 vs no-spiro)* | 28 (10.7%) | 0.31 (n/s) |
| *(p-value)* |  |  |  |  |  |  |
| Time to appearance (days) | 2.0 ± 0.8 | 1.6 ± 0.5<br>*(p=0.006 vs HA)*<br>*(n/a vs spiro)* | 2.5 ± 0.7<br>*(n/a vs spiro)* | 0<br>*(n/a vs no-spiro)* | 2.0 ± 0.8 | n/a |
| *(p-value)* |  |  |  |  |  |  |
| Duration (days) | 4.3 ± 2.1 | 4.3 ± 1.4<br>*(n/s (p=0.33) vs HA)*<br>*(n/a vs spiro)* | 4.2 ± 2.7<br>*(n/a vs spiro)* | 0<br>*(n/a vs no-spiro)* | 4.3 ± 2.1 | n/a |
| *(p-value)* |  |  |  |  |  |  |
| Lower back pain |  |  |  |  |  |  |
| Presence (%) | 14 (5.2%) | 10 (5.1%)<br>*(n/s (p=0.99) vs HA)*<br>*(n/s(p=0.81) vs spiro)* | 4 (6.0%)<br>*(n/s (p=0.78) vs spiro)* | 0<br>*(n/s (p=0.80) vs no-spiro)* | 14 (5.3%) | 0.96 (n/s) |
| *(p-value)* |  |  |  |  |  |  |
| Time to appearance (days) | 2.1 ± 0.7 | 1.8 ± 0.6<br>*(n/a vs HA)*<br>*(n/a vs spiro)* | 2.7 ± 0.4<br>*(n/a vs spiro)* | 0<br>*(n/a vs no-spiro)* | 2 ± 0.8 | n/a |
| *(p-value)* |  |  |  |  |  |  |
| Duration (days) | 3.3 ± 1.5 | 2.9 ± 1.2<br>*(n/a vs HA)*<br>*(n/a vs spiro)* | 4.2 ± 1.8<br>*(n/a vs spiro)* | 0<br>*(n/a vs no-spiro)* | 4.3 ± 2.1 | n/a |
| *(p-value)* |  |  |  |  |  |  |
| Diarrhea |  |  |  |  |  |  |
| Presence (%) | 65 (24.1%) | 41 (21.0%)<br>*(n/s (p=0.15) vs HA)*<br>*(ns (p=0.85) vs spiro)* | 22 (32.8%)<br>*(n/s (p=0.72) vs spiro)* | 2 (25.0%)<br>*(n/s (p=0.96) vs no-spiro)* | 63 (24.0%) | 0.35 (n/s) |
| *(p-value)* |  |  |  |  |  |  |
| Time to appearance (days) | 1.5 ± 0.8 | 1.4 ± 0.7<br>*(n/s (p=0.58) vs HA)*<br>*(n/a vs spiro)* | 1.5 ± 0.8<br>*(n/a vs spiro)* | 1.5 ± 0.5 (1;2)<br>*(n/a vs no-spiro)* | 1.5 ± 0.8 | n/a |
| *(p-value)* |  |  |  |  |  |  |
| Duration (days) | 3.2 ± 1.5 | 3.1 ± 1.3<br>*(n/s (p=0.41) vs HA)*<br>*(n/a vs spiro)* | 3.5 ± 1.7<br>*(n/a vs spiro)* | 1.5 ± 0.5 (1;2)<br>*(n/a vs no-spiro)* | 3.2 ± 1.5 | n/a |
| *(p-value)* |  |  |  |  |  |  |
| Nauseas |  |  |  |  |  |  |
| Presence (%) | 22 (8.1%) | 13 (6.7%)<br>*(n/s (p=0.65) vs HA)*<br>*(n/s (p=0.38) vs spiro)* | 7 (10.4%)<br>*(n/s (p=0.50) vs spiro)* | 2 (25.0%)<br>*(n/s (p=0.40) vs no-spiro)* | 20 (7.6%) | 0.63 (n/s) |
| *(p-value)* |  |  |  |  |  |  |
| Time to appearance (days) | 1.3 ± 0.5 | 1.2 ± 0.6<br>*(n/s (p=0.69) vs HA)*<br>*(n/a vs spiro)* | 1.3 ± 0.4<br>*(n/a vs spiro)* | 1.5 ± 0.5 (1;2)<br>*(n/a vs no-spiro)* | 1.2 ± 0.5 | n/a |
| *(p-value)* |  |  |  |  |  |  |
| Duration (days) | 1.7 ± 0.9 | 1.6 ± 0.7<br>*(n/s (p=0.53) vs HA)*<br>*(n/a vs spiro)* | 2.0 ± 1.1<br>*(n/a vs spiro)* | 1.0 ± 0.0 (1;1)<br>*(n/a vs no-spiro)* | 1.7 ± 0.9 | n/a |
| *(p-value)* |  |  |  |  |  |  |
| Vomiting |  |  |  |  |  |  |
| Presence (%)<br>(*p*-value) | 2 (0.7%) | 1<br>(*n/a* vs HA)<br>(*n/a* vs spiro) | 1 (1.5%)<br>(*n/a* vs spiro) | 0<br>(*n/a* vs no-spiro) | 2 (0.8%) | 1.0 (n/s) |
| Time to<br>appearance (days)<br>(*p*-value) | 2.0 ± 1.0 | 1<br>(*n/a* vs HA)<br>(*n/a* vs spiro) | 3<br>(*n/a* vs spiro) | 0<br>(*n/a* vs no-spiro) | (1;3) | n/a |
| Duration (days)<br>(*p*-value) | 2.0 ± 1.0 | 3<br>(*n/a* vs HA)<br>(*n/a* vs spiro) | 1<br>(*n/a* vs spiro) | 0<br>(*n/a* vs no-spiro) | (1;3) | n/a |
| **Abdominal pain** |  |  |  |  |  |  |
| Presence (%)<br>(*p*-value) | 21 (7.8%) | 13 (6.7%)<br>(*n/s* (*p*=0.52) vs HA)<br>(*n/s* (*p*=0.75) vs<br>spiro) | 8 (11.9%)<br>(*n/s*(*p*=0.58) vs<br>spiro) | 0<br>(*n/s* (*p*=0.70) vs no-spiro) | 21 (8.0%) | 0.75 |
| Time to<br>appearance (days)<br>(*p*-value) | 1.1 ± 0.3 | 1.1 ± 0.3<br>(*n/s* (*p*=1.0) vs HA)<br>(*n/a* vs spiro) | 1.1 ± 0.3<br>(*n/a* vs spiro) | 0<br>(*n/a* vs no-spiro) | 1.1 ± 0.3 | n/a |
| Duration (days)<br>(*p*-value) | 1.8 ± 0.7 | 1.7 ± 0.6<br>(*n/s* (*p*=0.23) vs HA)<br>(*n/a* vs spiro) | 2.1 ± 0.8<br>(*n/a* vs spiro) | 0<br>(*n/a* vs no-spiro) | 1.8 ± 0.7 | n/a |
| **Conjunctival**<br>**hyperemia** |  |  |  |  |  |  |
| Presence (%)<br>(*p*-value) | 22 (8.1%) | 9 (4.6%)<br>(*n/s*(*p*=0.071) vs HA)<br>(*n/s* (*p*=0.82) vs<br>spiro) | 13 (19.4%)<br>(*n/s* (*p*=0.37) vs<br>spiro) | 0<br>(*n/s* (*p*=0.69) vs no-spiro) | 22 (8.4%) | 0.18 |
| Time to<br>appearance (days)<br>(*p*-value) | 1.1 ± 0.5 | 1.2 ± 0.6<br>(*n/s* (*p*=0.88) vs HA)<br>(*n/a* vs spiro) | 1.1 ± 0.3<br>(*n/a* vs spiro) | 0<br>(*n/a* vs no-spiro) | 1.1 ± 0.5 | n/a |
| Duration (days)<br>(*p*-value) | 4.9 ± 2.6 | 4.2 ± 0.4<br>(*n/s* (*p*=0.54) vs HA)<br>(*n/a* vs spiro) | 5.5 ± 3.3<br>(*n/a* vs spiro) | 0<br>(*n/a* vs vs no-spiro) | 5.0 ± 2.6 | n/a |
| **Pre-orbital pain** |  |  |  |  |  |  |
| Presence (%)<br>(*p*-value) | 34 (12.6%) | 23 (11.8%)<br>(*n/s* (*p*=0.57) vs HA)<br>(*n/s* (*p*=0.57) vs<br>spiro) | 11 (16.4%)<br>(*n/s* (*p*=0.78) vs<br>spiro) | 0<br>(*n/s* (*p*=0.53) vs no-spiro) | 34 (13.0%) | 0.70 (n/s) |
| Time to<br>appearance (days)<br>(*p*-value) | 1.4 ± 0.6 | 1.3 ± 0.6<br>(*n/s* (*p*=0.27) vs HA)<br>(*n/a* vs spiro) | 1.5 ± 0.5<br>(*n/a* vs spiro) | 0<br>(*n/a* vs no-spiro) | 1.4 ± 0.6 | n/a |
| Duration (days)<br>(*p*-value) | 2.8 ± 1.3 | 2.4 ± 1.3<br>(***p*=0.016**) vs HA)<br>(*n/a* vs spiro) | 3.5 ± 1.1<br>(*n/a* vs spiro) | 0<br>(*n/a* vs no-spiro) | 2.8 ± 1.3 | n/a |
| **Dry eyes** |  |  |  |  |  |  |
| Presence (%)<br>(*p*-value) | 6 (2.2%) | 5 (2.6%)<br>(*n/s* (*p*=0.99) vs HA)<br>(*n/s* vs spiro) | 1 (1.5%)<br>(*n/s* vs spiro) | 0<br>(*n/s* vs no-spiro) | 6 (2.3%) | 0.98 (n/s) |
| Time to<br>appearance (days)<br>(*p*-value) | 1.4 ± 0.8 | 1.0 ± 0.0<br>(*n/a* vs HA)<br>(*n/a* vs spiro) | 3<br>(*n/a* vs spiro) | 0<br>(*n/a* vs no-spiro) | 1.4 ± 0.8 | n/a |
| Duration (days)<br>(*p*-value) | 6.2 ± 2.1 | 5.2 ± 1.1<br>(*n/a* vs HA)<br>(*n/a* vs spiro) | 10<br>(*n/a* vs spiro) | 0<br>(*n/a* vs no-spiro) | 6.2 ± 2.1 | n/a |
| **Dry mouth** |  |  |  |  |  |  |
| Presence (%)<br>(*p*-value) | 3 (1.1%) | 3 (1.5%)<br>(*n/s* (*p*=0.96) vs HA)<br>(*n/s* vs spiro) | 0<br>(*n/s* vs spiro) | 0<br>(*n/s* vs no-spiro) | 3 (1.1%) | 0.99 (n/s) |
| Time to<br>appearance (days) | 1.0 ± 0.0 | 1.0 ± 0.0<br>(*n/a* vs HA)<br>(*n/a* vs spiro) | 0<br>(*n/a* vs spiro) | 0<br>(*n/a* vs no-spiro) | 1.0 ± 0.0 | n/a |
| *(p-value)* |  |  |  |  |  |  |
| Duration (days) | 7.0 ± 0.0 | 7.0 ± 0.0 | 0 | 0 | 7.0 ± 0.0 | n/a |
| *(p-value)* |  | *(n/a vs HA)*<br>*(n/a vs spiro)* | *(n/a vs spiro)* | *(n/a vs no-spiro)* |  |  |

HA = hyperandrogenic; n/s = non-significant n/a = non-applicable

COVID-19 has shown the following sequence of symptoms, in a chronological order: 1. Earlier symptoms (< 2.0 days from the first symptom; in days): dizziness (*1*.*0 ± 0*.*2), abdominal pain* (*1*.*1 ± 0*.*3);* conjunctival hyperemia (*1*.*1 ± 0*.*5)*, nasal congestion or rhinorrhea (*1*.*2 ± 0*.*5)*, headache (*1*.*2 ± 0*.*5), dry cough* (*1*.*2 ± 0*.*5)*, myalgia (*1*.*2 ± 0*.*4)*, nauseas (*1*.*3 ± 0*.*5)*, weakness (*1*.*3 ± 0*.*5)*, “sinusitis” (*1*.*3 ± 0*.*5)*, hiporexia (*1*.*3 ± 0. 5)*, fatigue (*1*.*4 ± 0*.*7)*, pre-orbital pain (*1*.*4 ± 0*.*6), “sore throat”* (*1*.*4 ± 0*.*7)*, “feverish” (*1*.*5 ± 0*.*6*, fever (*1*.*5 ± 0*.*6)*, arthralgia (*1*.*5 ± 0*.*6) and diarrhea* (*1*.*5 ± 0*.*8);* and 2. Later symptoms (> 2.0 days since first symptom) include *upper back pain (2*.*0 ± 0*.*8); lower back pain (2*.*1 ± 0*.*7);* thoracic pain (*2*.*5 ± 1*.*0);* anosmia (*3*.*4 ± 1*.*3)*, ageusia (*3*.*4 ± 1*.*3)*, shortness of breath (*3*.*8 ± 1*.*2*).

Duration of symptoms during COVID-19 ranged from those with less than three days, including dizziness (*1*.*9 ± 1*.*0)*, abdominal pain (*1*.*8 ± 0*.*7)*, nauseas (*1*.*7 ± 0*.*9)*, fever (*2*.*3 ± 0*.*8);* “feverish” (*2*.*4 ± 1*.*2)*, shortness of breath (*2*.*5 ± 1*.*6)*, weakness (*2*.*7 ± 1*.*4)* and pre-orbital pain (*2*.*8 ± 1*.*3*); of intermediate duration (3 to 5 days), including arthralgia (*3*.*0 ± 2*.*4)*, diarrhea (*3*.*2 ± 1*.*5)*, myalgia (*3*.*3 ± 1*.*4)*, lower back pain (*3*.*3 ± 1*.*5)*, nasal congestion or rhinorrhea (*3*.*6 ± 1*.*8)*, upper back pain (*4*.*3 ± 2*.*1)*, thoracic pain (*4*.*7 ± 2*.*5)*, hyporexia (*4*.*8 ± 2*.*8)*, and conjunctival hyperemia (*4*.*9 ± 2*.*6)*; and those with prolonged duration (> 5 days), including dry cough (*5*.*4 ± 3*.*5)* “sore throat” (5.5 ± 1.9), headache (*5*.*7 ± 3*.*5)*, “sinusitis” (*6*.*0 ± 2*.*6), dry eyes* (*6*.*2 ± 2*.*1)*, ageusia (*7*.*0 ± 5*.*7), dry mouth* (*7*.*0 ± 0*.*0)*, fatigue (*7*.*2 ± 4*.*9) and* anosmia (*7*.*9 ± 6*.*2)*.

Similar time-to-appearance and duration: fever, “feverish”, nasal congestion or rhinorrhea, headache, shortness of breath, “sinusitis”, “sore throat”, dizziness, thoracic pain, lower back pain, diarrhea, nauseas, abdominal pain, pre-orbital pain, and dry eyes and mouth. Although conjunctival hyperemia was almost five times more present in HA than non-HA females (in terms of percentage), it did not reach statistical significance.

Anosmia and ageusia were almost absent in spironolactone users. Fewer non-HA than HA females were affected by anosmia, ageusia, dry cough, fatigue, weakness and hyporexia. Ageusia, weakness and myalgia lasted shorter in non-HA than HA. Time-to-appearance was shorter in non-HA for upper back pain.

As shown in Table 8, among proposed therapeutical options for COVID-19, besides azithromycin, which was given to all patients, nitazoxanide, hydroxychloroquine and ivermectin were prescribed in statistically equal proportions. There were also no differences between the percentage of additional drugs prescribed between groups (Table 9).

**Table 8.** Drugs prescribed for COVID-19.

| Medications<br>(+ azithromycin) | Overall<br>females<br>(n = 270) | Non-HA<br>females<br>(n = 195) | HA<br>females<br>(n = 67) | Spironolactone<br>Users (*spiro*)<br>(n = 8) | Non-spiro<br>(non-HA + HA)<br>(n = 262) | p-value<br>(overall) |
| --- | --- | --- | --- | --- | --- | --- |
| Hydroxychloroquine | 91 (33.7%) | 68 (34.8%) | 20 (29.8%) | 3 (37.5%) | 88 (33.6%) | 0.81 (n/s) |
| Nitazoxanide | 129 (47.8%) | 93 (47.7%) | 31 (43.3%) | 5 (62.5%) | 124 (47.3%) | 0.75 (n/s) |
| Ivermectin | 64 (23.7%) | 45 (23.1%) | 18 (26.9%) | 1 (12.5%) | 63 (24.0%) | 0.71 (n/s) |
| Spironolactone | 171 (63.3%**) | 124 (63.6%) | 47 (70.1%) | 0 | 171 (65.3%) | 0.42 (n/s) |

HA = hyperandrogenic; n/s = non-significant; n/a = non-applicable
\*\*Not including spironolactone users (n = 8)

**Table 9.** Additional drugs and supplements used to treat COVID-19.

| Additional<br>drugs or<br>supplements | Overall<br>females<br>(n = 270) | Non-HA<br>females<br>(n = 195) | HA<br>females<br>(n = 67) | Spironolactone<br>Users (*spiro*)<br>(n = 8) | Non-spiro (non-<br>+ HA) (n = 262) |
| --- | --- | --- | --- | --- | --- |
| Xa factor<br>inhibitors | 30 (11.1%) | 23 (11.8%) | 7 (10.4%) | 0 | 30 (11.4%) |
| Enoxaparin | 23 (8.5%) | 15 (7.7%) | 8 (11.9%) | 0 | 23 (8.8%) |
| Glucocorticoids | 34 (12.6%) | 24 (12.3%) | 10 (14.9%) | 0 | 34 (13.0%) |
| Vitamin C | 57 (21.1%) | 40 (20.5%) | 15 (22.4%) | 2 (25.0%) | 55 (21.0%) |
| Zinc | 68 (25.2%) | 48 (24.6%) | 17 (25.4%) | 3 (37.5%) | 65 (24.8%) |
| Vitamin D | 221 (81.8%) | 160 (82.0%) | 56 (83.6%) | 5 (62.5%) | 216 (82.4%) |

HA = hyperandrogenic; n/s = non-significant; n/a = non-applicable

### COVID-19 course and outcomes

Table 10 shows the average time-to-treat and the clinical and biochemical duration of COVID-19 manifestations. Time-to-treat, positive rtPCR, and duration of symptoms with and without anosmia and ageusia were all significantly lower in HA-spiro when compared to non-HA, HA, and overall non-users, even when only HA-spiro females with symptoms were included for the analysis. Conversely, while time-to-treat was similar, all other three outcomes were significantly shorter in non-HA compared to HA.

**Table 10.** COVID-19 clinical outcomes.

| Clinical clustering | Overall females (n = 270) | Non-HA females (n = 195) | HA females (n = 67) | Spiro lactone Users (*spiro*) (n = 8) | Non-spiro (non-HA + HA) (n = 262) | p-value (overall) |
| --- | --- | --- | --- | --- | --- | --- |
| **Time-to-treat**<br>(*p*-value) | 3.0 ± 1.8 | 3.0 ± 1.9<br>(*n*/s (*p*=0.11) vs HA)<br>(*p*<0.007 vs *spiro*) | 3.4 ± 1.4<br>(*p*=0.0001 vs *spiro*) | 0.5 ± 1.1 (Median = 0; 95%CI = 0.7)<br>(*p*=0.0003 vs no-*spiro*)<br>(*p* = 0.0028 vs no-*spiro* after adjustment for symptomatic patients only) | 3.1 ± 1.8 | 0.0004 |
| **Duration of positive rtPCR (days)**<br>(*p*-value) | 13.9 ± 6.1 | 13.5 ± 5.7<br>(*p*=0.033 vs vs HA)<br>(*p*=0.025 vs *spiro*) | 15.7 ± 6.7<br>(*p*=0.0005 vs *spiro*) | 8.7 ± 3.0<br>(*p*=0.015 vs no-*spiro*)<br>(*p* = 0.015 vs no-*spiro* after adjustment for symptomatic patients only) | 14.0 ± 6.1 | 0.0057 |
| **Remission not including anosmia (days)**<br>(without (*p*-value)) | 5.8 ± 4.7 | 5.5 ± 4.7<br>(*p*=0.0003 vs vs HA)<br>(*p*=0.0008 vs *spiro*)<br>(*p* = 0.015 vs *spiro* after adjustment for symptomatic patients only) | 7.2 ± 4.5<br>(*p*<0.0001 vs *spiro*)<br>(*p*=0.0002 vs *spiro* after adjustment for only symptomatic patients) | 1.4 ± 1.4 (Median = 1; 95%CI = 1.0)<br>(*p*=0.0003 vs no-*spiro*)<br>(*p* = 0.0046 vs vs no-*spiro* after adjustment for symptomatic patients only) | 6.0 ± 4.7 | < 0.0001<br>(*p* < 0.0001 after adjustment for only symptomatic patients) |
| **Remission including anosmia (days)**<br>(without (*p*-value)) | 9.6 ± 7.3 | 9.0 ± 7.1<br>(*p*=0.0001 vs HA)<br>(*p*=0.0004 vs *spiro*)<br>(*p*=0.0046 vs *spiro* after adjustment for symptomatic patients only) | 12.4 ± 7.0<br>(*p*<0.0001 vs *spiro*)<br>(*p*<0.0001 vs *spiro* after adjustment for only symptomatic patients) | 1.7 ± 2.0 (Median = 1; 95%CI = 0.3)<br>(*p*=0.0001 vs no-*spiro*)<br>(*p* = 0.0013 vs no-*spiro* after adjustment for symptomatic patients only) | 9.9 ± 7.3 | < 0.0001<br>(<0.0001 after adjustment for only symptomatic patients) |

HA = hyperandrogenic; n/s = non-significant; n/a = non-applicable

Table 11 details the clinical course of COVID-19. Although the peak of symptoms is apparently higher in the HA group, it did not reach statistical significance when compared to non-HA due to the wide variability. Conversely, the recovery process seems to be slower in this group, as evidenced by the largest differences between HA and non-HA, and between HA and HA-spiro in Days 2 to 7, which coincides with the early response to proposed pharmacological treatments. HA-spiro group had fewer symptoms than non-spironolactone users in Days -3 to -1 and Day 7, as well as than HA females in Days -3 to -1 and Days 2 to 7.

**Table 11.** COVID-19 course.

| Clinical evolution | Overall females (n = 270) | Non-HA females (n = 195) | HA females (n = 67) | Spironolactone Users (*spiro*) (n = 8) | Non-spiro (non-HA + HA) (n = 262) | p-value (overall) |
| --- | --- | --- | --- | --- | --- | --- |
| **Days -7 to -4**<br>(*p*-value) | 80.2 ± 36.3 (Median = 100; 95%CI = 4.3)<br>(75.9% asymptomatic) | 78.1 ± 37.9 (Median = 100; 95%CI = 5.3)<br>[144 (73.8%)<br>asymptomatic]<br>(n/s (p=0.41) vs HA)<br>(n/s (p=0.21) vs spiro) | 84.1 ± 32.6 (Median = 100; 95%CI = 7.8) [53 (79.1%)<br>asymptomatic]<br>(n/s (p=0.34) vs spiro) | 100.0 ± 0 (Median = 100; 95%CI = 0) [8 (100%)<br>asymptomatic]<br>(n/s (p=0.23) vs no-spiro) | 79.6 ± 36.7 (Median = 100; 95%CI = 4.4)<br>[197 (75.2%)<br>asymptomatic] | 0.35 (n/s) |
| **Days -3 to -1**<br>(*p*-value) | 34.5 ± 40.0 (Median = 10; 95%CI = 4.8)<br>(24.1% asymptomatic) | 36.2 ± 40.8 (Median = 10; 95%CI = 5.7) [49 (24.0%) asymptomatic]<br>(n/s (p=0.41) vs AGA vs HA)<br>(p=0.007 vs spiro) | 23.4 ± 31.0 (Median = 10; 95%CI = 7.4) [9 (13.4%)<br>asymptomatic]<br>(p=0.003 vs spiro) | 87.5 ± 33.1 (Median = 100; 95%CI = 22.9) [7 (87.5%)<br>asymptomatic (p=0.005 vs no-spiro) | 32.9 ± 39.0 (Median = 10; 95%CI = 4.7)<br>[58 (22.1%)<br>asymptomatic] | 0.014 |
| **Day 0**<br>(*p*-value) | 32.0 ± 35.0 (Median = 20; 95%CI = 4.2)<br>(12.6% asymptomatic) | 35.0 ± 36.6 (Median = 20; 95%CI = 5.1) [28 (14.3%) asymptomatic]<br>(p=0.045) vs AGA vs HA)<br>(n/s (p=0.29) vs spiro) | 20.1 ± 23.1 (Median = 15; 95%CI = 5.5) [2 (3.0%)<br>asymptomatic]<br>(n/s (p=0.16) vs spiro) | 57.5 ± 46.3 (Median = 80; 95%CI = 32.1) [4 (50.0%)<br>asymptomatic]<br>(n/s (p=0.25) vs no-spiro) | 31.2 ± 34.3 (Median = 20; 95%CI = 4.2)<br>[30 (11.4%)<br>asymptomatic] | 0.072 (n/s) |
| **Day 1**<br>(*p*-value) | 55.9 ± 33.6 (Median = 60; 95%CI = 4.0)<br>(12.6% asymptomatic) | 59.9 ± 33.0 (Median = 70; 95%CI = 4.6) [28 (14.3%) asymptomatic]<br>(p=0.0002 vs HA)<br>(n/s (p<0.30) vs spiro) | 43.0 ± 30.3 (Median = 40; 95%CI = 7.3) [2 (3.0%)<br>asymptomatic] (n/s (p=0.12) vs spiro) | 65.0 ± 44.5 (Median = 97.5; 95%CI = 30.8) [4 (50.0%)<br>asymptomatic] (n/s (p=0.23) vs no-spiro) | 55.6 ± 33.1 (Median = 60; 95%CI = 4.0)<br>[30 (11.4%)<br>asymptomatic] | 0.0008 |
| **Day 2**<br>(*p*-value) | 75.1 ± 27.0 (Median = 90; 95%CI = 3.2)<br>(20.4% asymptomatic) | 78.0 ± 26.0 (Median = 90; 95%CI = 3.6) [46 (23.6%) asymptomatic]<br>(p<0.0001 vs HA)<br>(p=0.0051) vs spiro) | 65.4 ± 26.8 (Median = 70; 95%CI = 6.4) [4 (6.0%)<br>asymptomatic]<br>(p=0.0007 vs spiro) | 83.75 ± 32.7 (Median = 100; 95%CI = 22.6) [5 (62.5%)<br>asymptomatic]<br>(p=0.044 vs no-spiro) | 74.8 ± 26.8 (Median = 82.5; 95%CI = 3.2)<br>[50 (19.1%)<br>asymptomatic] | < 0.0001 |
| **Day 3**<br>(*p*-value) | 85.7 ± 22.5 (Median = 95; 95%CI = 2.7)<br>(36.3% asymptomatic) | 87.0 ± 22.8 (Median = 95; 95%CI = 3.2) [81 (41.5%) asymptomatic]<br>(p=0.0007 vs HA)<br>(p<0.0001 vs spiro) | 80.3 ± 21.6 (Median = 90; 95%CI = 5.2) [9 (13.4%)<br>asymptomatic]<br>(p=0.0001 vs spiro) | 100.0 ± 0 (Median = 100; 95%CI = 0) [8 (100%)<br>asymptomatic]<br>(p=0.0016 vs no-spiro) | 85.3 ± 22.7 (Median = 90; 95%CI = 2.7)<br>[90 (34.3%)<br>asymptomatic] | < 0.0001 |
| **Day 7**<br>(*p*-value) | 94.0 ± 14.3 (Median = 100; 95%CI = 1.7)<br>(66.3% asymptomatic) | 94.1 ± 15.5 (Median = 100; 95%CI = 2.2) [137 (70.2%)<br>asymptomatic] (p=0.023 vs HA)<br>(n/s (p=0.15) vs spiro) | 93.0 ± 11.3 (Median = 100; 95%CI = 2.7) [34 (50.7%)<br>asymptomatic]<br>(p=0.023 vs spiro) | 100.0 ± 0 (Median = 100; 95%CI = 0) [8 (100%)<br>asymptomatic]<br>(n/s (p=0.094) vs no-spiro) | 93.8 ± 14.5 (Median = 100; 95%CI = 1.8)<br>[171 (65.3%)<br>asymptomatic] | 0.018 |
| **Day 14**<br>(*p*-value) | 98.5 ± 7.6 (Median = 100; 95%CI = 0.9)<br>(90.0% asymptomatic) | 98.5 ± 8.4 (Median = 100; 95%CI = 1.2) [178 (91.3%)<br>asymptomatic]<br>(n/s (p=0.45) vs HA)<br>(n/s (p=0.67) vs spiro) | 98.3 ± 5.2 (Median = 100; 95%CI = 1.2) [57 (85.1%)<br>asymptomatic]<br>(n/s (p=0.49) vs spiro) | 100.0 ± 0 (Median = 100; 95%CI = 0) [8 (100%)<br>asymptomatic]<br>(n/s (p=0.62) vs no-spiro) | 98.4 ± 7.7 (Median = 100; 95%CI = 0.9)<br>[235 (89.7%)<br>asymptomatic] | 0.67 (n/s) |
| **Day 21**<br>(*p*-value) | 99.4 ± 6.3 (Median = 100; 95%CI = 0.7)<br>(97.4% asymptomatic) | 99.4 ± 7.2 (Median = 100; 95%CI = 1.0) [191 (97.9%)<br>asymptomatic]<br>(n/s vs HA)<br>(n/s vs spiro) | 99.5 ± 2.7 (Median = 100; 95%CI = 0.7) [64 (95.2%)<br>asymptomatic]<br>(n/s vs spiro) | 100.0 ± 0 (Median = 100; 95%CI = 0) [8 (100%)<br>asymptomatic]<br>(n/s vs no-spiro) | 99.4 ± 6.3 (Median = 100; 95%CI = 0.9)<br>[235 (89.7%)<br>asymptomatic] | 1.0 (n/s) |
| **Day 30**<br>(*p*-value) | 99.8 ± 3.0 (Median = 100; 95%CI = 0.4)<br>(98.5% asymptomatic) | 99.7 ± 3.6 (Median = 100; 95%CI = 0.5) [191 (97.9%)<br>asymptomatic]<br>(n/s vs HA)<br>(n/s vs spiro) | 100.0 ± 0 (Median = 100; 95%CI = 0.0) [67 (100%)<br>asymptomatic]<br>(n/s vs spiro) | 100.0 ± 0 (Median = 100; 95%CI = 0) [8 (100%)<br>asymptomatic]<br>(n/s vs no-spiro) | 99.8 ± 3.1 (Median = 100; 95%CI = 0.8)<br>[255 (98.5%)<br>asymptomatic] | 1.0 (n/s) |
| **Day 60**<br>(*p*-value) | 100.0 ± 0 (Median = 100; 95%CI = 0) (100% asymptomatic) | 100.0 ± 0 (Median = 100; 95%CI = 0) [195 (100%) asymptomatic] (n/s vs HA) (n/s vs spiro) | 100.0 ± 0 (Median = 100; 95%CI = 0.0) [67 (100%) asymptomatic] (n/s vs spiro) | 100.0 ± 0 (Median = 100; 95%CI = 0) [8 (100%) asymptomatic] (n/s vs no-spiro) | 100.0 ± 0.0 (Median = 100; 95%CI = 0.0) [262 (100%) asymptomatic] | 1.0 (n/s) |

HA = hyperandrogenic; n/s = non-significant; n/a = non-applicable; CI = confidence interval

As in Table 12, which depicts the World Health Organization (WHO) COVID Ordinal Outcomes in the present population, HA females had significantly higher scores than non-HA and HA-spiro groups in Days 0 and 7, and became similar after Day 14. None of the patients presents as Scores 3 to 5 throughout the disease.

**Table 12.**
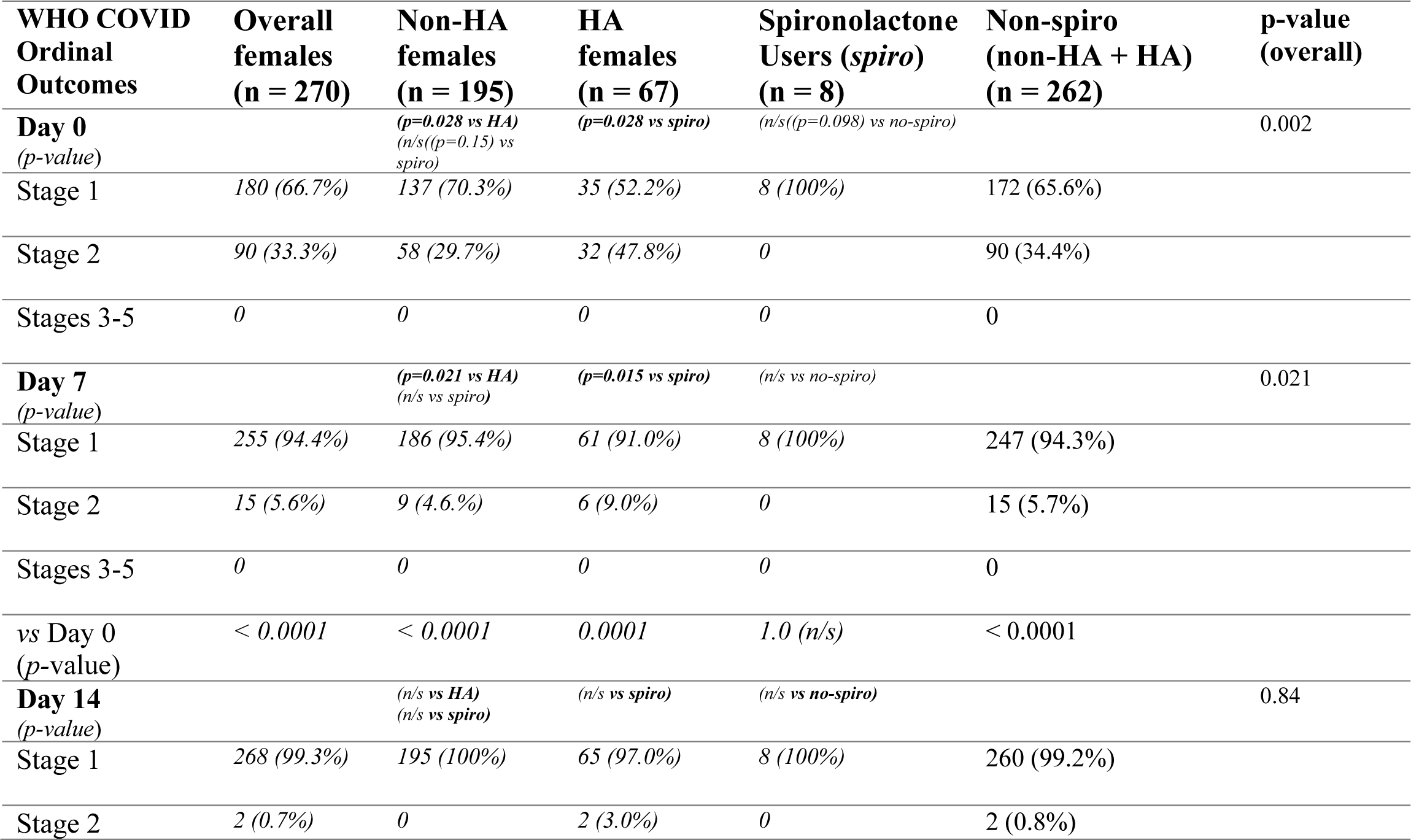
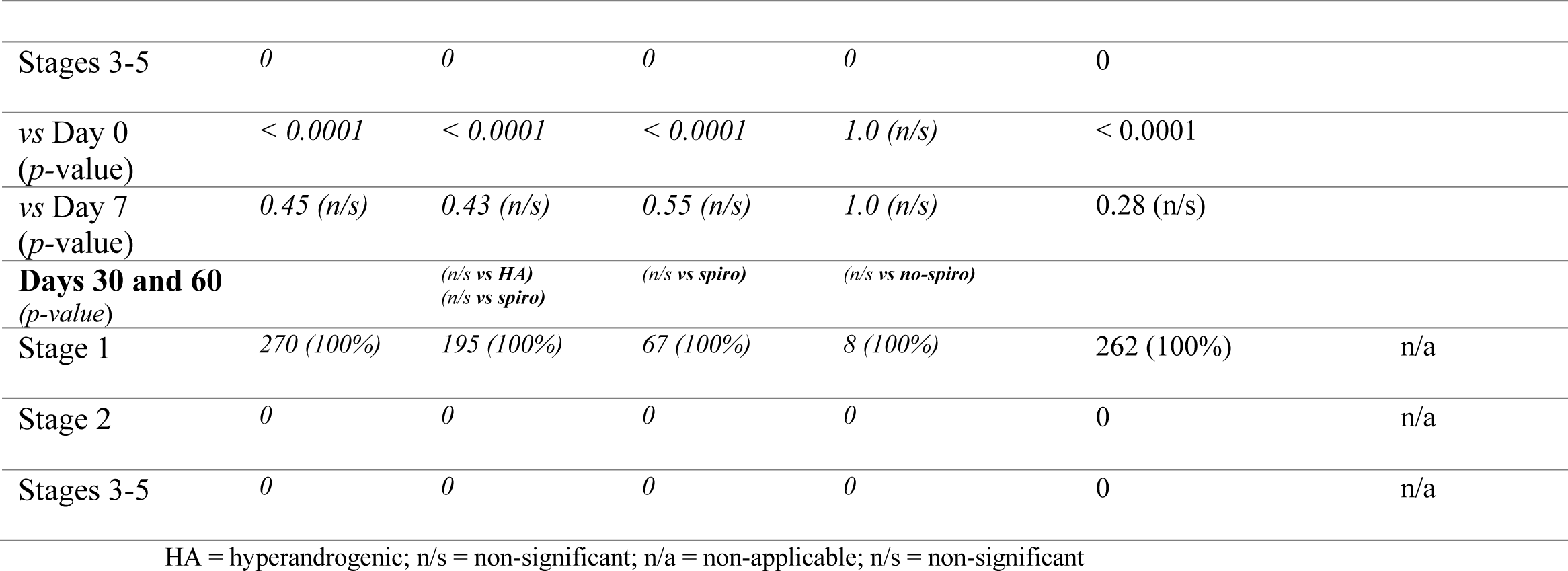
WHO COVID-19 Ordinal Outcomes.

The loss of capacity to perform everyday activities is described in Table 13. None of the spironolactone users had any level of loss of everyday activities. Conversely, HA were significantly more compromised when compared to non-HA and HA-spiro females (p = 0.004 and 0.028, respectively), by the time that proposed treatments started (Day 0). This was mitigated in the following days (Days 3, 7, 14 and 30). Except for spiro-HA (not affected at any time), Day 3 had significant improvement compared to Day 0.

**Table 13.**
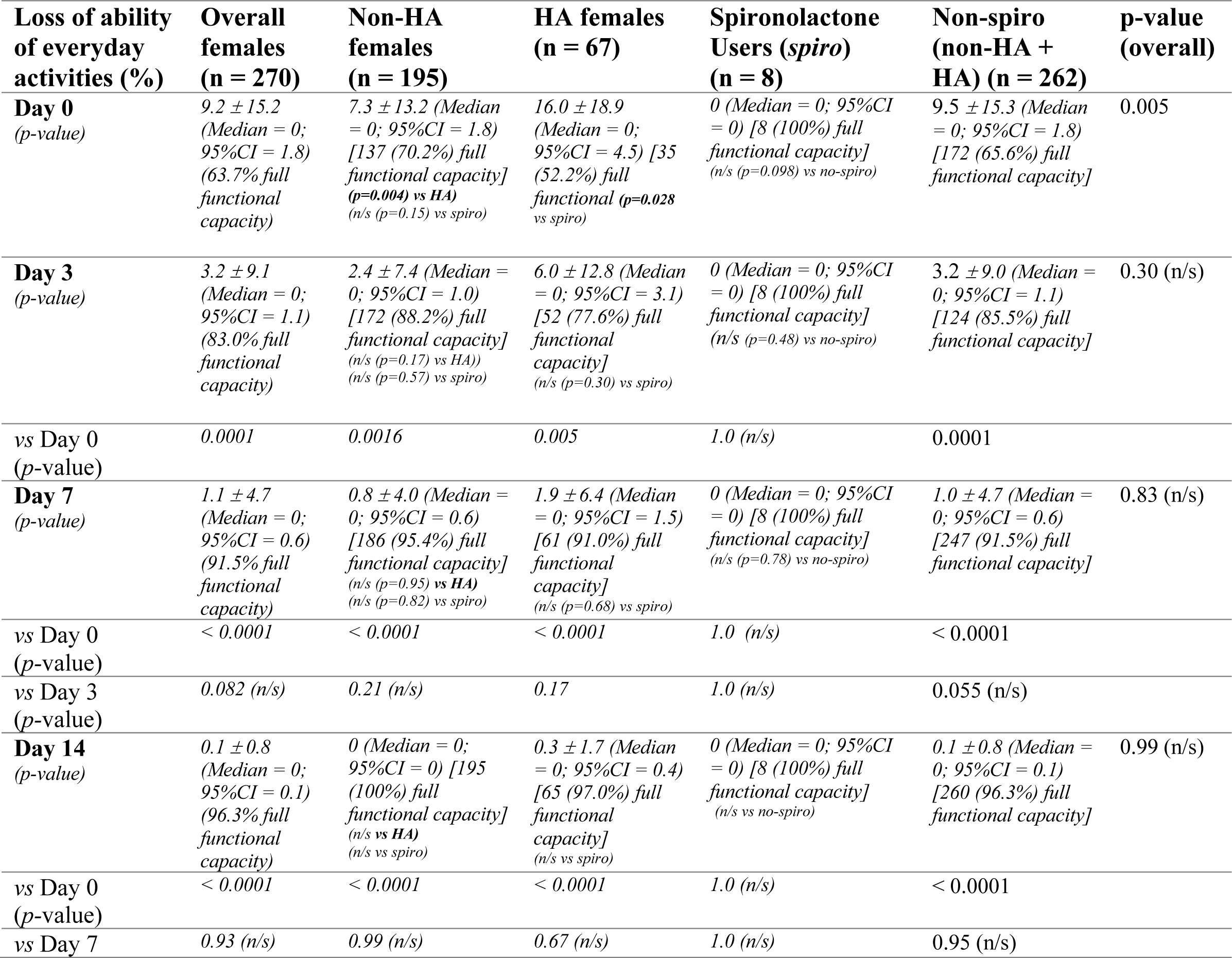
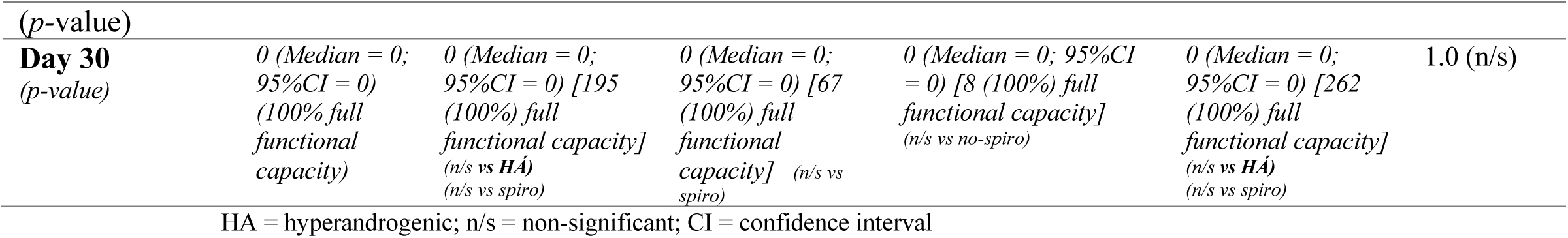
Loss of ability to perform everyday activities due to COVID-19.

Table 14 estimates the percentage of lung affected in COVID-19 in Days 0, 7, 14 and 30 through chest computerized tomography (CT) scan. Because none of HA-spiro reported severe symptoms, none of these patients underwent CT. Non-HA had significantly less lungs affected than HA females, except for Day 30. None of these two groups had significant improvement

**Table 14.** Chest CT scan.

| Chest CT scan (% of lungs affected) | Overall females (n = 270) | Non-HA females (n = 195) | HA females (n = 67) | Spirolactone Users (spiro) (n = 8) | Non-spiro (non-HA + HA) (n = 262) | p-value (overall) |
| --- | --- | --- | --- | --- | --- | --- |
| **Day 0**<br>(p-value) | 14.7 ± 14.8 (Median = 10; 95%CI = 1.8) [32 (34.4%) normal lungs of 93 patients that performed chest CT scan in day 0] | 11.8 ± 13.3 (Median = 10; 95%CI = 1.9) [25 (41.0%) normal lungs of 61 patients that performed chest CT scan in day 0] (p=0.015 vs HA) (n/a vs spiro) | 20.3 ± 15.7 (Median = 25; 95%CI = 3.8) [7 (21.89%) normal lungs of 32 patients that performed chest CT scan in day 0] (n/a vs spiro) | n/a (no patients performed chest CT scan) | (= overall) | n/a |
| **Day 7**<br>(p-value) | 15.0 ± 14.6 (Median = 10; 95%CI = 1.7) [20 (31.25%) normal lungs of 64 patients that performed chest CT scan in day 7] | 11.9 ± 12.9 (Median = 10; 95%CI = 1.8) [15 (38.4%) normal lungs of 39 patients that performed chest CT scan in day 7] (p=0.053 vs HA) (n/a vs spiro) | 19.8 ± 15.8 (Median = 10; 95%CI = 3.8) [5 (20.0%) normal lungs of 25 patients that performed chest CT scan in day 7] (n/a vs spiro) | n/a | (= overall) | n/a |
| vs Day 0<br>(p-value) | 0.85 (n/s) | 0.89 (n/s) | 0.92 (n/s) | n/a |  |  |
| **Day 14**<br>(p-value) | 11.2 ± 11.7 (Median = 10; 95%CI = 1.4) [23 (39.0%) normal lungs of 59 patients that performed chest CT scan in day 14] | 7.3 ± 9.0 (Median = 0; 95%CI = 1.3) [19 (51.3%) normal lungs of 37 patients that performed chest CT scan in day 14] (p=0.003 vs HA) (n/a vs spiro) | 17.7 ± 12.8 (Median = 17.5; 95%CI = 3.0) [4 (18.1%) normal lungs of 22 patients that performed chest CT scan in day 14] (n/a vs spiro) | n/a (no patients performed chest CT scan) | (= overall) | n/a |
| vs Day 0<br>(*p*-value) | 0.25 (n/s) | 0.18 (n/s) | 0.62 (n/s) | n/a |  |  |
| vs Day 7<br>(*p*-value) | 0.21 (n/s) | 0.17 (n/s) | 0.75 (n/s) | n/a |  |  |
| **Day 30**<br>(*p*-value) | 10.9 ± 10.3 (Median = 17.5; 95%CI = 1.4) [6 (37.5%) normal lungs of 16 patients that performed chest CT scan in day 30] | 9.6 ± 10.0 (Median = 10; 95%CI = 1.1) [5 (41.7%) normal lungs of 12 patients that performed chest CT scan in day 30] (n/s (*p*=0.32) vs HA) (n/a vs spiro) | 15.0 ± 10.6 (Median = 17.5; 95%CI = 2.5) [1 (25.0%) normal lungs of 4 patients that performed chest CT scan in day 30] (n/a vs spiro) | n/a (no patients performed chest CT scan) | (= overall) | n/a |
| vs Day 0<br>(*p*-value) | 0.49 (n/s) | 0.78 (n/s) | 0.44 (n/s) | 1.0 (n/s) |  |  |
| vs Day 7<br>(*p*-value) | 0.44 (n/s) | 0.72 (n/s) | 0.53 (n/s) | 1.0 (n/s) |  |  |
| vs Day 14<br>(*p*-value) | 0.95 (n/s) | 0.52 (n/s) | 0.67 (n/s) | 1.0 (n/s) |  |  |

HA = hyperandrogenic; n/s = non-significant; CI = confidence interval

Table 15 summarizes the scores of COVID-19 progression and severity, and complications related to the disease. None of the 270 patients enrolled in the present study presented any complication for COVID-19 throughout the treatment.

**Table 15.** COVID-19 progression outcomes.

| **Disease progression outcomes** | **Overall females (n = 270)** | **Non-HA females (n = 195)** | **HA females (n = 67)** | **Spirolactone Users (*spiro*) (n = 8)** | **p-value (overall)** |
| --- | --- | --- | --- | --- | --- |
| **Brescia COVID-19 Respiratory Severity Scale (0-4)** | 0 | 0 | 0 | 0 | 1.00 (n/s) |
| **Hospitalization** | 0 | 0 | 0 | 0 | 1.00 (n/s) |
| **Mechanical ventilation** | 0 | 0 | 0 | 0 | 1.00 (n/s) |
| **Noradrenaline/dopamine** | 0 | 0 | 0 | 0 | 1.00 (n/s) |
| **Death** | 0 | 0 | 0 | 0 | 1.00 (n/s) |

HA = hyperandrogenic; n/s = non-significant; CI = confidence interval

## Discussion

### The importance of an early diagnosis of COVID-19

The present prospective observational study aimed to characterize patients upon the diagnosis of COVID-19. Oppositely to the prevailing recommendations to suspect for COVID-19 only in the presence of fever of shortness of breath, we detected COVID-19 guided by any symptom, related or supposedly unrelated to the disease. Another key modification was the active questions, since patients tend to avoid reporting symptoms that they do not believe to be due to COVID-19. The majority of patients had at least one complain that would not be reported if not actively questioned.

Researches have largely focused approaches for stages two and three, while assumed that none of proposed therapies for earlier stages would be effective (‘wait and see’). Culturally, unlike bacterial infections, specific antiviral treatments tend to be avoided. Except for chronic and relapsing viruses, antivirals have been studied at an irregular basis, and their clinical use have long been avoided. Moreover, drugs for other diseases that might disclose direct or indirect antiviral activity have been underestimated.

More important than the culture of non-viral treatment that surrounds the medical field, is that the lack of specific symptoms and the usual presence of uncommon clinical presentations, that likely precluded the majority of the patients from an actual early diagnosis.

The change in the paradigm for detection of COVID-19 towards more sensitive and active search for COVID-19 may have driven the better outcomes observed in the present study. We consider imperative that policies for COVID-19 diagnosis change

### The patient with COVID-19: characterization of the patients and the disease

Baseline characteristics were significantly different between non-HA and HA probably because HA females tend to be shorter and present higher BMI. The differences between age is understandable when one considers that the phenotypical expression of HA tends to occur earlier in lifetime. Furthermore, recognition of female HA (except for dramatic cases, as beard females) has only occurred in the last decades, and was underdiagnosed until short time ago.

Differences in BMI between HA and non-HA was relatively lower than differences in age, and obesity was similarly present between groups. Conversely, even being significantly younger, HA females presented more severe manifestations of COVID-19 than non-HA

While none of the prevailing chronic diseases presented any difference between groups, metformin and oral contraceptives were more used in HA than non-HA females, possibly due to the fact the PCOS is the most common underlying condition that leads to HA.

In a similar manner of what has been observed in males, symptoms in females could be easily clustered (23-26). At least two symptoms of each cluster were together within a same patient in more than 80% of patients. The clusters of anosmia-ageusia predominance, dengue fever-like, URTI-like, and GI infection-like clinical presentation have been detected, in addition to those with unspecific symptoms, or with more than one cluster. The remarkable URTI-like clinical symptoms among HA females may be due to enhanced infectivity that has been shown to occur under hyperandrogenic states.

The similar number of patients treated with hydroxychloroquine, nitazoxanide and ivermectin and the lack of major differences at least in terms of COVID-19 complications shows that the choice for a specific medication to be added to azithromycin is not imperative. Also, although preliminary hypotheses and data demonstrating the important role of azithromycin as an adjuvant therapy against SARS-CoV-2, whether this drug plays an add-on benefit effect is uncertain, and remains unanswered, since we did not compare with *versus* without its use.

Correspondingly, additional drugs and supplements were given according to an individualized clinical evaluation, and were used in similar percentages between groups. The use of anticoagulant, specially Xa factor inhibitors and enoxaparin, were based on the risk for thrombosis and development of other complications, since COVID-19 is a pro-thrombotic state *per se*. Vitamin D, and at slightly lesser extent, zinc and vitamin C, were prescribed to the majority of treated patients. Hence, whether these additional prescriptions helped to improve outcomes cannot be concluded from the present study.

### COVID-19 outcomes

All early COVID-19 related outcomes, including duration of positive rtPCR-SAR-CoV-2 and symptoms with or without anosmia and ageusia were markedly lower in HA-spiro when compared to non-HA, HA, and overall non-users. However, time-to-treatment was equally lower in HA-spiro females. Hence, whether the shorter and milder clinical presentation in COVID-19 among spironolactone users is due to earlier beginning of treatment is unknown, although COVID-19 disease course remained better in the HA-spiro group after adjustments for non-asymptomatic patients only.

Unlike HA-spiro, since time-to-treat and treatment options were similar between non-HA and HA females, obesity prevalence was similar, and HA were younger than non-HA, prolonged clinical and viral duration among HA females may only be explained by the hyperandrogenism.

Although spironolactone users had more asymptomatic presentation than other groups, when HA-spiro patients presented symptoms, these symptoms presented similar time-to-appearance and duration than non-HA females. The great majority of the symptoms occurred in similar percentages and presented similar durations for all groups. Increased fatigue, dry cough and anosmia, and increased and prolonged weakness, myalgia and ageusia found in HA females should also be explained by hyperandrogenism, in an analogical manner than male AGA (5,6,14,16,17), due to enhanced expression of TMPRSS-2.

Differences in disease course were more evident between Days 0 and 7 in HA females, as this population presented slower speed of improvement compared to other groups, as evidenced by three methods to quantify this speed (Tables 11 to 13), which means that HA females were shown to improve, but took longer than other groups. Despite the slower symptom regression in HA, all groups showed increased recovery rate after initiation of any of the proposed therapies.

The importance of measuring loss of capacity to perform everyday activities lays on the fact that infected patients remain isolated for approximately 14 days, and needs to be able to perform the basic hygiene and self-care activities during this period. Although HA females were more compromised than non-HA and HA-spiro females, even this group was only slightly affected, and lasted for less than three days.

Unlike rapid clinical recovery, radiology did not show any improvement during the first 30 days after treatment initiation. However, there is a major bias that those that underwent more than one chest CT were those that persisted with complains or were already compromised. Hence, radiological improvement may be less prolonged that what has been shown in the present study.

Criteria employed to measure COVID-19 severity, ilike WHO COVID Scale and Brescia Respiratory Scale, revealed to be inaccurate in the present analysis, since none of the patients progressed to respiratory complications, hospitalization, and other outcomes. Instead, more sensitive scales were better to detect differences.

Oppositely to non-spironolactone HA females, HA that use spironolactone had markedly better clinical outcomes related to COVID-19. Additionally, the small number of patients with COVID-19 taking spironolactone may reflect its potential preventive effect, despite the lack of any other data to support this hypothesis. Spironolactone has been proposed to exert multiple beneficial roles in COVID-19, including increase of circulating-to-attached angiotensin converting enzyme 2 (ACE2) ratio, increased angiotensin receptor (AT) 2-to-AT1 ratio, decreased TMPRSS2 activity, as well as antiviral and anti-inflammatory activities (20-22), and its promising roles have been reinforced by the present analysis.

### Post-COVID syndrome?

The importance of unveiling potential effective treatments for COVID-19 not only lays on preventing hospitalization, mechanical ventilation, and death, but also preventing long-term, post-cure persisting symptoms, termed as “post-COVID syndrome”, that has shown to be present in at least 20 to 30% of patients, and includes autoimmune, mental, psychiatric, muscular, respiratory and gastrointestinal disorders, and persisted fatigue, resembling chronic fatigue syndrome (CFS), unrelated to any specific biochemical marker to date (27,28). Hence, whenever a pharmacological intervention study is conducted, researchers should not only consider acute, but also chronic complications as targets to evaluate efficacy.

### The effect of specific treatments on COVID-19: comparison with existing literature

When duration and severity of clinical symptoms, disease duration, and related complications are compared with the extensive reports on the literature, differences are undisputable, particularly for clinical outcomes (10-12). At least in the present population of females, differences were sufficiently overwhelming to justify the lack of need for placebo-control trials, from an ethical perspective. However, open label RCTs and other prospective observational studies are mandatory to confirm the clear findings of the present study, when compared to the expected outcomes for COVID-19 in females.

### Limitations

As a prospective observational study conducted prior to a double-blind placebo-control RCT, aiming to better determine the selection process and parameters to be evaluated, as well as define the most plausible pharmacological approach between hydroxychloroquine, nitazoxanide, ivermectin, or none. As per the study deisgn, the lack of a placebo group initially hampers from conclusive findings. However, this has been overcome by the evident differences when compared to the well-established COVID-19 course and outcomes, which disallows us from performing the RCT as a full placebo-control.

The replication of a highly sensitive case-detection guidance that include the occurrence of absolutely any symptom as being suspected for COVID-19 may find barriers that may preclude from a successful approach, including: 1. Lack of general and medical education regarding the unspecific pattern of COVID-19 clinical presentation; 2. Self-judgement of not being affected by COVID-19; 3. Inability to correlate non-obvious symptoms with COVID-19, losing the window of opportunity; and 4. Cultural focus on the severe patient, neglecting those are possibly preventable from COVID-19 complication. Although this may not necessarily occur with all patients, the larger number of patients treated early, the better clinical outcomes should be.

### Final discussion

Since COVID-19 is an extensive, diffuse, and largely unclarified disease, a thorough medical evaluation that encompasses questions for all organs and systems should be performed at a regular basis, in clinical practice, not only restricted to researching purposes.

The early detection associated with any drug combination among azithromycin, hydroxychloroquine, nitazoxanide and ivermectin yielded notable improvements in terms of course of COVID-19, when compared to literature.

Among females, presenting HA may be an additional risk for COVID-19 severity, as they presented to be slightly more affected and had a prolonged recovery process when compared to non-HA and spironolactone users. The chronic use of spironolactone in a daily dose 100mg or more in HA females not only seemed to mitigate, but also reduce risks to below those found in non-HA. This has shown to have similar relative risks when compared to the presence of AGA and use of dutasteride among males, respectively.

## Conclusion

A sensitive, early detection of COVID-19 followed by a pharmaceutical approach with a drug combination between azithromycin in all cases, associated with hydroxychloroquine, nitazoxanide or ivermectin demonstrated unequivocal differences when compared to the extensively described natural disease course when pharmacological treatments potentially effective for COVID-19 is not provided, particularly if detected until three days after the beginning of symptoms.

When compared to non-hyperandrogenic (non-HA), HA females had a more severe and prolonged clinical presentation, even though risk of development of COVID-19 related complications remained unmeaningful among all groups. Spironolactone mitigated any additional risk due to HA.

## Data Availability

Full raw data is available at a data repository (https://osf.io/cm4f8/).

https://osf.io/cm4f8/

## References

1. Guan W, Ni Z, Hu Y, et al. Clinical characteristics of coronavirus disease 2019 in China. N Engl J Med 2020; Feb 28.

2. Hajifathalian K, Kumar S, Newberry C, et al. Obesity is associated with worse outcomes in COVID-19: Analysis of Early Data From New York City [published online ahead of print, 2020 May 29]. Obesity (Silver Spring). 2020;10.1002/oby.22923.

3. Kalligeros M, Shehadeh F, Mylona EK, et al. Association of Obesity with Disease Severity among Patients with COVID-19 [published online ahead of print, 2020 Apr 30]. Obesity (Silver Spring). 2020; 10.1002/oby.22859.

4. Palaiodimos L, Kokkinidis DG, Li W, et al. Severe obesity, increasing age and male sex are independently associated with worse in-hospital outcomes, and higher in-hospital mortality, in a cohort of patients with COVID-19 in the Bronx, New York [published online ahead of print, 2020 May 16]. Metabolism. 2020;108:154262.

5. Cavalcanti AB, Zampieri FG, Rosa RG, Azevedo LCP, Veiga VC, Avezum A, Damiani LP, Marcadenti A, Kawano-Dourado L, Lisboa T, Junqueira DLM, de Barros E Silva PGM, Tramujas L, Abreu-Silva EO, Laranjeira LN, Soares AT, Echenique LS, Pereira AJ, Freitas FGR, Gebara OCE, Dantas VCS, Furtado RHM, Milan EP, Golin NA, Cardoso FF, Maia IS, Hoffmann Filho CR, Kormann APM, Amazonas RB, Bocchi de Oliveira MF, Serpa-Neto A, Falavigna M, Lopes RD, Machado FR, Berwanger O; Coalition Covid-19 Brazil I Investigators. Hydroxychloroquine with or without Azithromycin in Mild-to-Moderate Covid-19. N Engl J Med. 2020 Jul 23:NEJMoa2019014. doi:10.1056/NEJMoa2019014.

6. Zhou F, Yu T, Du R, et al. Clinical course and risk factors for mortality of adult inpatients with COVID-19 in Wuhan, China: a retrospective cohort study [published online ahead of print, 2020 Mar 11] [published correction appears in Lancet. 2020 Mar 12;:]. Lancet. 2020;S0140-6736(20)30566-3.

7. Lauer SA, Grantz KH, Bi Q, et al. The Incubation Period of Coronavirus Disease 2019 (COVID-19) From Publicly Reported Confirmed Cases: Estimation and Application [published online ahead of print, 2020 Mar 10]. Ann Intern Med. 2020;10. 7326/M20-0504.

8. Wu C, Chen X, Cai Y, et al. Risk Factors Associated With Acute Respiratory Distress Syndrome and Death in Patients With Coronavirus Disease 2019 Pneumonia in Wuhan, China [published online ahead of print, 2020 Mar 13]. JAMA Intern Med. 2020; 10.1001/jamainternmed.2020.0994.

9. Wu C, Chen X, Cai Y, et al. Risk Factors Associated With Acute Respiratory Distress Syndrome and Death in Patients With Coronavirus Disease 2019 Pneumonia in Wuhan, China [published online ahead of print, 2020 Mar 13]. JAMA Intern Med. 2020; 10.1001/jamainternmed.2020.0994.

10. https://www.who.int/teams/health-care-readiness-clinical-unit/covid-19/data-platform (Last accessed September 29th, 2020)

11. https://www.cdc.gov/coronavirus/2019-ncov/hcp/clinical-guidance-management-patients.html (Last accessed September 29th, 2020)

12. Stokes EK, Zambrano LD, Anderson KN, et al. Coronavirus Disease 2019 Case Surveillance - United States, January 22-May 30, 2020. MMWR Morb Mortal Wkly Rep 2020; 69:759.

13. Cadegiani, F.A. Repurposing existing drugs for COVID-19: an endocrinology perspective. BMC Endocr Disord 20, 149 (2020).https://doi.org/10.1186/s12902-020-00626-0

14. Pascarella G, Strumia A, Piliego C, Bruno F, Del Buono R, Costa F, Scarlata S, Agrò FE. COVID-19 diagnosis and management: a comprehensive review. J Intern Med. 2020 Aug;288(2):192–206. doi:10.1111/joim.13091.

15. Goren A, Vano-Galvan S, Wambier CG, et al. A preliminary observation: male pattern hair loss among hospitalized COVID-19 patients in Spain - A potential clue to the role of androgens in COVID-19 severity [published online ahead of print, 2020 Apr 16]. J Cosmet Dermatol. 2020;10. 1111/jocd.13443.

16. Goren A, McCoy J, Wambier CG, et al. What does androgenetic alopecia have to do with COVID-19? An insight into a potential new therapy [published online ahead of print, 2020 Apr 1]. Dermatol Ther. 2020;e13365.

17. McCoy J, Wambier CG, Herrera S, Vaño-Galván S, Gioia F, Comeche B, Ron R, Serrano-Villar S, Iwasiow RM, Tayeb MA, Cadegiani FA, Mesinkovska NA, Shapiro J, Sinclair R, Goren A. Androgen Receptor Genetic Variant Predicts COVID-19 Disease Severity: A Prospective Longitudinal Study of Hospitalized COVID-19 Male Patients. J Eur Acad Dermatol Venereol. 2020 Sep 25.

18. Kragholm K, Andersen MP, Gerds TA, et al. Association between male sex and outcomes of Coronavirus Disease 2019 (Covid-19) - a Danish nationwide, register-based study. Clin Infect Dis 2020.

19. Goren A, Wambier CG, Herrera S, McCoy J, Vaño-Galván S, Gioia F, Comeche B, Ron R, Serrano-Villar S, Ramos PM, Cadegiani FA, Kovacevic M, Tosti A, Shapiro J, Sinclair R. Anti-androgens may protect against severe COVID-19 outcomes: results from a prospective cohort study of 77 hospitalized men. J Eur Acad Dermatol Venereol. 2020 Sep 25. doi: 10.1111/jdv.16953.

20. Cadegiani FA. Can spironolactone be used to prevent COVID-19-induced acute respiratory distress syndrome in patients with hypertension? Am J Physiol Endocrinol Metab. 2020 May 1;318(5):E587–E588. doi: 10.1152/ajpendo.00136.2020.

21. Cadegiani FA, Wambier CG, Goren A. Spironolactone: An Anti-androgenic and Anti-hypertensive Drug That May Provide Protection Against the Novel Coronavirus (SARS-CoV-2) Induced Acute Respiratory Distress Syndrome (ARDS) in COVID-19. Front Med (Lausanne). 2020 Jul 28;7:453. doi: 10.3389/fmed.2020.00453.

22. Cadegiani FA, Goren A, Wambier CG. Spironolactone may provide protection from SARS-CoV-2: Targeting androgens, angiotensin converting enzyme 2 (ACE2), and renin-angiotensin-aldosterone system (RAAS). Med Hypotheses. 2020 Jul 16;143:110112. doi: 10.1016/j.mehy.2020.110112.

23. https://www.uptodate.com/contents/coronavirus-disease-2019-covid-19-clinicalfeatures?topicRef=126981&source=see_link (Last accessed September 29th, 2020)

24. Kim GU, Kim MJ, Ra SH, Lee J, Bae S, Jung J, Kim SH. Clinical characteristics of asymptomatic and symptomatic patients with mild COVID-19. Clin Microbiol Infect. 2020 Jul;26(7):948.e1-948.e3. doi: 10.1016/j.cmi.2020.04.040.

25. Lechien JR, Chiesa-Estomba CM, De Siati DR et al. Olfactory and gustatory dysfunctions as a clinical presentation of mild-to-moderate forms of the coronavirus disease (COVID-19): a multicenter European study. Eur Arch Otorhinolaryngol. 2020 Aug;277(8):2251–2261. doi: 10.1007/s00405-020-05965-1.

26. Tong JY, Wong A, Zhu D, Fastenberg JH, Tham T. The Prevalence of Olfactory and Gustatory Dysfunction in COVID-19 Patients: A Systematic Review and Meta-analysis. Otolaryngol Head Neck Surg. 2020 Jul;163(1):3–11. doi: 10.1177/0194599820926473.

27. Novak P. Post COVID-19 syndrome associated with orthostatic cerebral hypoperfusion syndrome, small fiber neuropathy and benefit of immunotherapy: a case report. eNeurologicalSci. 2020 Dec;21:100276. doi: 10.1016/j.ensci.2020.100276.

28. Clauw DJ, Häuser W, Cohen SP, Fitzcharles MA. Considering the potential for an increase in chronic pain after the COVID-19 pandemic. Pain. 2020 Aug;161(8):1694–1697. doi: 10.1097/j.pain.0000000000001950.

